# The induction of dissociative states: A meta-analysis

**DOI:** 10.1101/2024.09.09.24313338

**Authors:** Benjamin Brake, Lillian Wieder, Natasha Hughes, Ivonne Saravia Lalinde, Danielle Marr, Dali Geagea, Susannah Pick, Antje A. T. S. Reinders, Sunjeev K. Kamboj, Trevor Thompson, Devin B. Terhune

## Abstract

**Objective:** Dissociative states, characterised by discontinuities in awareness and perception, occur in a diverse array of psychiatric disorders and contexts. Dissociative states have been experimentally modelled in the laboratory through various induction methods but relatively little is known about the efficacy and comparability of different experimental methods.

**Methods:** This meta-analysis quantified dissociative states, as indexed by a standardised instrument (*Clinician Administered Dissociative States Scale*), at baseline in varied diagnostic categories and in response to different experimental induction methods (psychological techniques and pharmacological agents) in both clinical and non-clinical samples. Primary outcomes were state dissociation effect sizes (Hedges’s *g*) (PROSPERO registration CRD42022384886).

**Results:** 2,214 papers were screened, yielding 123 eligible articles and 155 effect sizes comprising 6,692 individuals. High levels of baseline state dissociation were observed in multiple diagnostic categories relative to controls, with the largest effects found in the dissociative and complex subtypes of post-traumatic stress disorder (PTSD-DC). In controlled experiments, induced state dissociation was most pronounced in response to mirror-gazing and multiple pharmacological agents with effects exceeding baseline state dissociation in PTSD-DC in ketamine and cannabis. The effect sizes were characterised by pronounced heterogeneity but were not reliably associated with methodological features of the original studies.

**Conclusions:** Elevated state dissociation is present in multiple diagnostic categories and comparable or higher levels can be reliably induced in controlled experiments using psychological techniques and pharmacological agents. These results demonstrate the efficacy of several methods for experimentally modelling dissociation and have implications for measuring adverse events and predicting outcomes in clinical interventions involving pharmacological agents.

## Introduction

Dissociation comprises a constellation of symptoms characterised by discontinuities in awareness, volition, and perception (1, 2). These experiences range from episodes of depersonalisation and derealisation, encompassing feelings of detachment from emotional or bodily states, and/or one’s environment, respectively, to distortions in control, identity and memory. Dissociation is increasingly recognised as a transdiagnostic symptom prevalent in a wide variety of psychiatric conditions (2). Elevated levels of dissociation may also serve as a salient marker of clinical outcomes including a higher burden of illness (3), poorer quality of life (4), more pronounced symptomatology (5–7), and poorer treatment outcomes (8).

The clinical significance of dissociation underscores the need for controlled research on these symptoms but there exists no consensus experimental model of dissociation. Psychological techniques range from those that induce dissociative states through modulation of awareness and perception (e.g., mirror-gazing) or exposure to stressors (9). Multiple pharmacological agents have been shown to trigger dissociation, particularly those that function as N-methyl-D-aspartate receptor (NMDAR) antagonists, such as ketamine and nitrous oxide (N_2_O) (10). To our knowledge, there has not yet been any attempt to quantitatively synthesise and contrast these different induction effects, nor to compare them against baseline dissociative states in diagnostic categories.

A robust experimental model of dissociative states will offer novel opportunities for identifying neurophysiological and neurochemical markers of dissociative states, elucidating the impact of dissociation on other symptoms (e.g., hallucinations), and could inform both the diagnosis and treatment of a range of psychiatric conditions (10, 11). Moreover, as NMDAR antagonists and serotonergic psychedelics are used or proposed as mainstream antidepressants (12), studying their dissociative effects might aid in advancing understanding of treatment-related adverse events (13) and treatment outcomes (14), which often covary with dissociative responses.

This meta-analysis sought to fill outstanding gaps in current knowledge regarding the experimental induction of dissociative states and their comparability to baseline dissociation in diagnostic categories. As in other meta-analyses (2), we sought to increase uniformity of comparisons within and across categories and thus restricted our analyses to studies that measured dissociative states using the *Clinician-Administered Dissociative States Scale* (CADSS) (15), the most widely used measure of state dissociation (16). Our primary aims were to quantitatively synthesise and compare baseline state dissociation effects in different diagnostic categories and in induced state dissociation effects in response to different psychological techniques and pharmacological agents. Our secondary aims were to explore the factors that moderate the magnitude of state dissociation effects within and across categories.

## Method

This pre-registered study (<u>t.ly/I-ppg</u>) was conducted under the updated PRISMA 2020 guidelines (17).

### Eligibility criteria

The inclusion criteria were: English language; full article in a peer-reviewed journal; participants aged 18 or older; inclusion of descriptive statistics and sample sizes for the CADSS in a diagnostic group and non-clinical control group or in an experimental and control condition. Exclusion criteria included: reviews, abstracts, dissertations, or case studies; data overlapping with included studies; use of a dissociation-attenuating agent; and CADSS completion after an extended period (>12h).

### Search strategy

In October 2022, two researchers (BB and LW) independently searched MEDLINE, PubMed, PsycINFO, and Embase using terms relating to the CADSS (Supplementary Materials). The search was limited to studies published since 1998, the CADSS’s initial publication year. All eligible studies were integrated into a database using Covidence ® (Veritas Health Innovation, Melbourne, Australia; available at www.covidence.org). The search was repeated in June 2023 and March 2024, yielding 6 and 4 additional studies, respectively.

### Study selection

Two independent raters (BB, DG, NH, DM, ISL, LW) independently screened and assessed all studies for eligibility using a two-stage procedure. First, they screened titles and abstracts, rejecting articles not meeting eligibility criteria. Then, they reviewed the remaining papers to finalize the study list. A third reviewer (DBT) resolved discrepancies at either stage. If eligible articles lacked sufficient CADSS data, corresponding authors were contacted via email (up to three attempts over three months).

### Data extraction

Data extraction was performed by two raters (BB, DG, NH, DM, ISL, LW). The primary outcomes extracted were CADSS scores (15) in a target condition/group and a control condition/group. Secondary outcomes included CADSS subscale scores and correlations between trait dissociation scores and CADSS scores. Both raters independently extracted and coded data using a pre-piloted extraction form in Covidence, covering: study details (authors, title, journal, publication date, country); demographics (sample size, gender distributions, age, education, ethnicity); study design (repeated-measures, between-groups, mixed-model); category (diagnostic group, psychological technique, pharmacological agent); CADSS information (administrator [clinician/experimenter v. self-report], mode of administration [in person or remote], version [number of items], number of measurement timepoints, subscales, language); trait dissociation measure; clinical study methods (diagnosis, diagnostic criteria, diagnostic method, comorbidities, control type [healthy or clinical], clinical control diagnosis); pharmacological study methods (CADSS measurement times, drug class, dose, administration method and duration, concurrent drug-use information, active/inert placebo information); psychological technique (method, control condition/group information); other methodological details (counterbalancing, inclusion of suggestion for dissociation); descriptive statistics for CADSS scores (total and subscales in all conditions); and correlations between trait dissociation and CADSS scores. If descriptive statistics were not reported, they were extracted from figures using WebPlotDigitizer (v. 4.6; https://automeris.io/) when possible. Discrepancies were resolved with a third reviewer and sometimes a fourth. Overall, there was 91% agreement between raters (range: 85-98%).

### Methodological quality

Two raters independently assessed the quality of each study using a 15-item scale (Supplementary Materials) concerning study objectives, participant recruitment, demographic data, inclusion/exclusion criteria, clarity of procedure, blinding, pre-registration, and relative matching of groups/conditions. The items, adapted from a previous meta-analysis (18), were based on Cochrane criteria and PRISMA recommendations (19). Each item was categorically rated (0=criterion not met, 1=met), and a percentage met total was computed for each study; DBT resolved discrepancies. There was 90% agreement between raters (range: 63%-100%; mean kappa=.80; range: .25-1).

### Meta-analysis and meta-regression

Descriptive statistics (*M*s, *SD*s, and *n*s, or other suitable statistics) were used to compute Hedges’s *g*s (and *SE*s) for inclusion in random effects meta-analyses when there were three or more effect sizes per category (Supplementary Methods). Multiple effect sizes were extracted and included in meta-analyses for specific categories only when data from distinct samples were reported (see Supplementary Table 2). For studies that reported multiple timepoints for the same participants (e.g., ketamine studies), we included only the effect size corresponding to the peak response. Some categories had an insufficient number of effect sizes and were thus consolidated into higher-order categories. Categories included different diagnostic categories, psychological techniques, and pharmacological agents. For each category, we computed standardised mean differences (*SMD*s) and 95% confidence intervals (CIs) after outlier removal (studentized residuals > |3.3| (20)). Although analysis of raw CADSS scores could permit a clearer comparison of effect sizes across categories, this approach was not feasible due to variations in the versions of the CADSS used by different author groups, which differ in both the included items and of scores (16). *SMD*s were coded such that positive values reflect greater state dissociation (CADSS score) in the reference category than a control.

Meta-analyses were supplemented with prediction intervals (PIs) when *k*≥5 (21, 22); PIs estimate the distribution of the effect in a future individual study with similar features. Heterogeneity of effect sizes was computed using *I*^2^ and τ^2^ where values exceeding 50% and 10%, respectively, reflect moderate or greater heterogeneity. Publication bias was evaluated using funnel plots of *SMD*s against *SE*s and Egger’s bias test, where *p*<.05 reflects asymmetry (23); we computed revised *SMD*s correcting for asymmetry using the trim-and-fill method (24). Moderators of effect sizes were assessed using meta-regression analyses where there were at least five effects sizes in each category and at least 10 effect sizes within a category, respectively. Multiple pre-registered analyses were not performed due to insufficient number of effect sizes or insufficient information in original papers (Supplementary Materials). Analyses were performed in JASP (v. 0.18.3, 2014; JASP Team, the Netherlands), Jamovi (v. 2.3.26.0, the Jamovi project), and MATLAB (v. 2023a, MathWorks, Natick, Massachusetts, USA).

## Results

### Study inclusion and characteristics

A PRISMA diagram showing study selection is presented in Supplementary Figure 1. 123 papers met inclusion criteria, yielding 155 effect sizes (*n*=6,629) that could be included in our main analysis categories (see Supplementary Results for exclusions). After excluding 9 outliers, the effect sizes included controlled comparisons of diagnostic categories (*k*=32, *n*=1,729), psychological techniques (*k*=50, *n*=2,400), or pharmacological agents (*k*=64, *n*=2,563) (Table 1). The largest categories (*k*s≥10) included PTSD, mirror-gazing, trauma stimuli exposure, and ketamine. Methodological quality ratings and study details can be found in Supplementary Table 2.

**Table 1.** Results of meta-analyses of state dissociation effects (CADSS scores in reference vs. control) as a function of diagnostic category, psychological technique, and pharmacological agent.

| Category | *k* | *n* | *SMD* | 95% CI | *PIs* | *Z* | *p* | *I*<sup>2</sup> (%) | *T*<sup>2</sup> | FPAp | Outliers |
| --- | --- | --- | --- | --- | --- | --- | --- | --- | --- | --- | --- |
| **Diagnostic categories** |  |  |  |  |  |  |  |  |  |  |  |
| PTSD-DC | 7 | 443 | 1.34 | [0.86, 1.82] | [-0.23, 2.91] | 5.44 | <.001 | 78.77 | .31 | .013 | 0 |
| PTSD | 12 | 644 | 0.94 | [0.65, 1.23] | [0.04, 1.84] | 6.42 | <.001 | 61.66 | .14 | .030 | 0 |
| MDD | 6 | 338 | 0.89 | [0.43, 1.35] | [-0.63, 2.41] | 3.82 | <.001 | 75.27 | .24 | .77 | 0 |
| SZ | 3 | 146 | 0.86 | [0.51, 1.21] | [-1.49, 3.21] | 4.83 | <.001 | 0 | 0 | .34 | 0 |
| FND | 4 | 158 | 0.59 | [-0.17, 1.35] | [-2.84, 4.02] | 1.52 | .13 | 80.59 | .48 | .086 | 0 |
| **Psychological techniques** |  |  |  |  |  |  |  |  |  |  |  |
| Mirror-gazing | 12 | 392 | 0.94 | [0.52, 1.35] | [-0.63, 2.51] | 4.44 | <.001 | 86.43 | .45 | <.001 | 0 |
| Military training | 9 | 639 | 0.77 | [0.52, 1.02] | [-0.07, 1.61] | 6.10 | <.001 | 80.1 | .11 | .37 | 1 |
| Sleep deprivation | 3 | 110 | 0.56 | [0.26, 0.86] | [-2.69, 3.81] | 3.69 | <.001 | 52.2 | .04 | .063 | 1 |
| Trauma stimuli | 18 | 902 | 0.50 | [0.36, 0.64] | [-0.01, 1.00] | 6.94 | <.001 | 63.68 | .05 | .15 | 1 |
| Complementary methods | 3 | 232 | 0.41 | [0.14, 0.67] | [-1.76, 2.58] | 2.99 | .003 | 22.29 | .01 | .23 | 0 |
| Negative affect stimuli | 5 | 125 | 0.16 | [-0.02, 0.34] | [-0.15, 0.47] | 1.79 | .074 | 0 | 0 | .20 | 1 |
| **Pharmacological agents** |  |  |  |  |  |  |  |  |  |  |  |
| Ketamine | 47 | 1,579 | 1.51 | [1.23, 1.80] | [0.17, 2.85] | 13.70 | <.001 | 83.65 | .42 | <.001 | 4 |
| Cannabis | 4 | 139 | 1.40 | [0.96, 1.83] | [-1.39, 4.19] | 6.30 | <.001 | 72.76 | .37 | .002 | 0 |
| N<sub>2</sub>O | 3 | 129 | 1.16 | [0.91, 1.41] | [-0.53, 2.85] | 9.00 | <.001 | 0 | 0 | .42 | 0 |
| Psychedelics | 4 | 68 | 1.16 | [0.66, 1.67] | [-0.91, 3.23] | 4.50 | <.001 | 63.03 | .16 | .34 | 1 |
| Esketamine | 6 | 648 | 0.94 | [0.55, 1.33] | [-0.28, 2.16] | 4.77 | <.001 | 78.87 | .15 | .001 | 0 |

Notes. *k* = number of included effect sizes (after removal of outliers); *N* = sample size; *SMD* = standardized mean difference; *PIs* = prediction intervals; *I*<sup>2</sup> = heterogeneity statistic; *T*<sup>2</sup> = heterogeneity statistic; FPAp = funnel plot asymmetry *p*-value; outliers = number of outliers removed; FND = functional neurological disorder; MDD = major depressive disorder; N<sub>2</sub>O = nitrous oxide; PTSD = post-traumatic stress disorder; PTSD-DC = post-traumatic stress disorder - dissociative subtypes and complex subtypes; SZ = schizophrenia.

**Figure 1.**
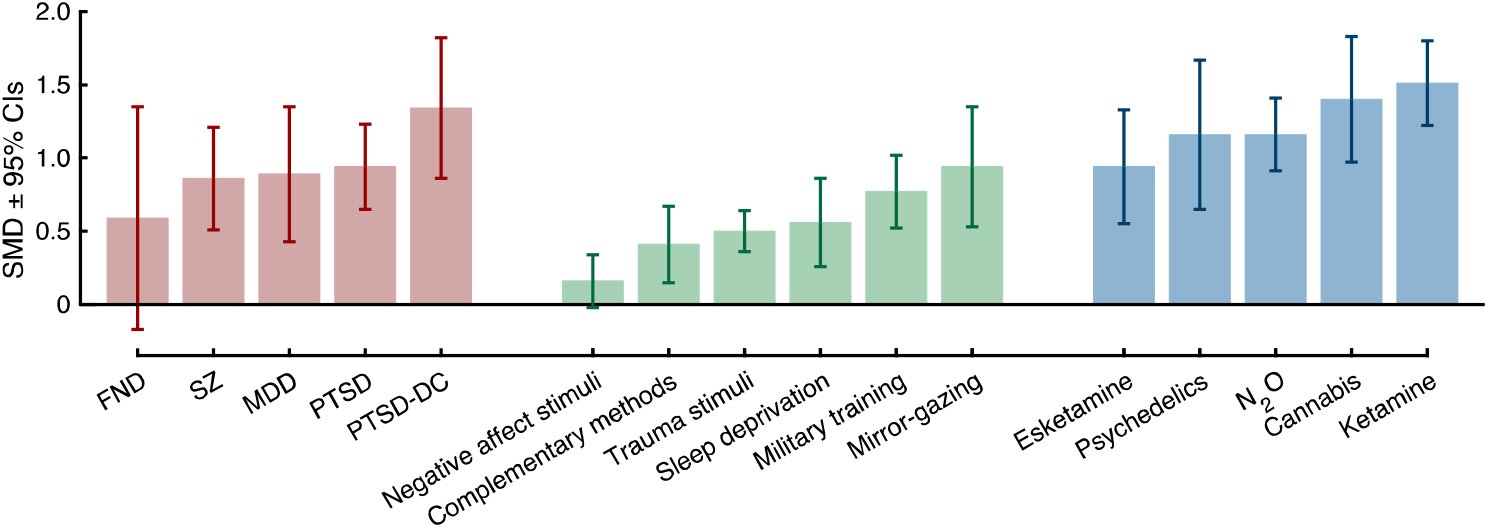
Results of meta-analyses of state dissociation (CADSS scores) at baseline in diagnostic categories (red), in response to psychological interventions (green), and in response to pharmacological agents (blue). CADSS = clinician administered dissociative states scale; SMD=Standardised Mean Difference; FND = functional neurological disorder; SZ = schizophrenia; MDD = major depressive disorder; PTSD = post-traumatic stress disorder; PTSD-DC = PTSD dissociative and complex subtypes; N_2_O = nitrous oxide.

### Meta-analyses of controlled comparisons of state dissociation in diagnostic categories

The magnitude of state dissociation at baseline was examined in five diagnostic groups in order to provide a reference point for effect sizes for induced dissociative states (Table 1, Figure 1). Dissociative states were significantly greater than non-clinical controls in all groups except functional neurological disorder (FND). Baseline state dissociation was most pronounced in patients meeting criteria for the dissociative and complex subtypes of PTSD (PTSD-DC) (*SMD*=1.34; Figure 2), followed by weaker, yet still large, effects in PTSD, MDD, and schizophrenia, which displayed comparable effect sizes (analyses of CADSS subscales were not possible due to an insufficient number of studies) (for forest plots, see Supplementary Materials). Prediction intervals were only significant in PTSD whereas moderate-to-large heterogeneity was observed in all groups except schizophrenia.

**Figure 2.**
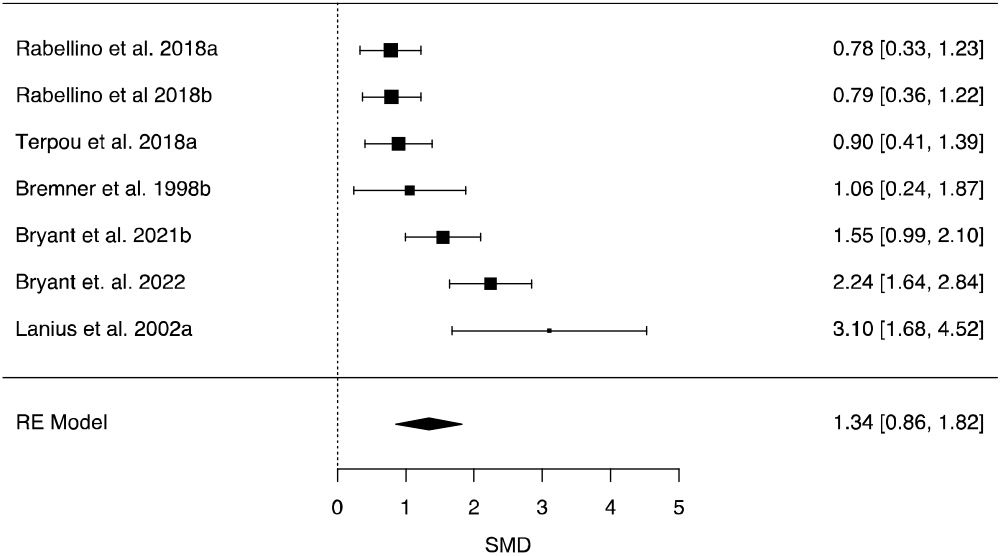
Forest plot of *Standardised Mean Differences* (*SMD*s; with 95% Confidence Intervals [CIs]) of baseline state dissociation (CADSS scores) in post-traumatic stress disorder dissociative and complex subtypes (PTSD-DC) relative to controls. Marker sizes reflect study weights with smaller markers denoting smaller study weights.

### Meta-analyses of psychological techniques for the induction of dissociative states

All induction methods significantly increased state dissociation except negative affect stimuli exposure (Table 1, Figure 1). Mirror-gazing was associated with a large effect size (*SMD*=0.94; see Figure 3), followed by a large effect for military training whereas sleep deprivation, trauma stimuli exposure, and complementary methods elicited moderate effects (for forest plots, see Supplementary Materials). Prediction intervals were non-significant for all methods. Moderate-to-large heterogeneity in effect sizes was observed for mirror-gazing, military training, and trauma stimuli exposure. Further analyses suggested that the effects of mirror-gazing were most pronounced for derealisation (Supplementary Table 3).

**Figure 3.**
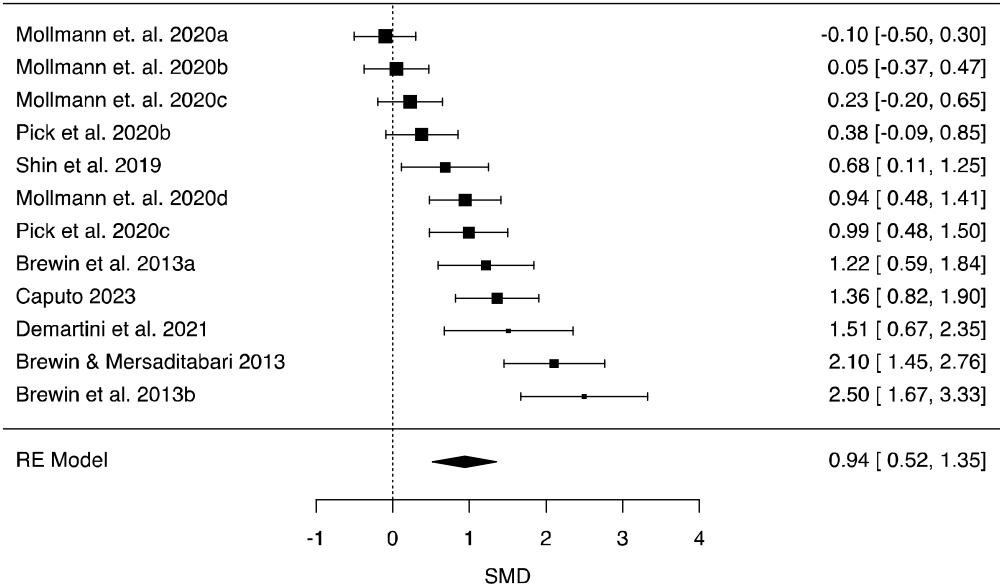
Forest plot of *Standardised Mean Differences* (*SMD*s; with 95% Confidence Intervals [CIs]) of induced state dissociation (CADSS scores) in response to mirror-gazing relative to a control condition. Marker sizes reflect study weights with smaller markers denoting smaller study weights.

### Meta-analyses of pharmacological induction of dissociative states

The analyses of pharmacological agents revealed that all agents reliably induced dissociative states with large effect sizes (Table 1, Figure 1, for forest plots, see Supplementary Materials). The largest effect sizes were observed for ketamine (Figure 4) and cannabis (*SMD*s>1.35), with slightly weaker effects for N_2_O and psychedelics (*SMD*s=1.16) and the weakest, albeit still large, effect for esketamine. Prediction intervals were significant only for ketamine and moderate-to-large heterogeneity in effect sizes was observed for all agents except N_2_O. Analyses of subdimensions of dissociation yielded comparable results although the effects were generally larger for derealisation across all agents (see Supplementary Table 3). Further analyses suggested that the dissociative effects were most pronounced during the first 30 minutes post-infusion (see Supplementary Table 4).

**Figure 4.**
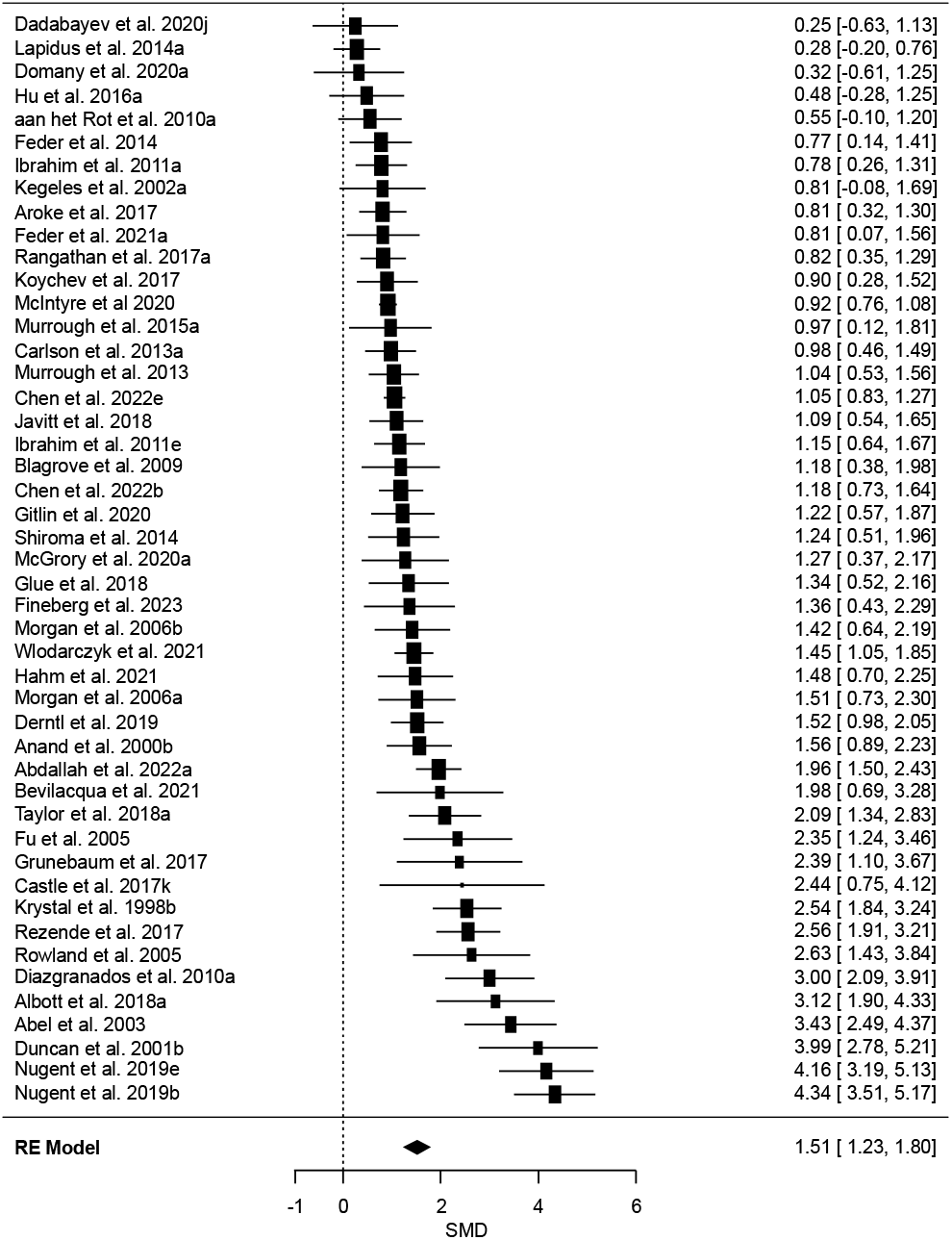
Forest plot of *Standardised Mean Differences* (*SMDs*; with 95% Confidence Intervals [CIs]) of induced state dissociation (CADSS scores) in response to ketamine relative to a control condition. Marker sizes reflect study weights with smaller markers denoting smaller study weights.

### Publication bias

Among diagnostic categories and psychological techniques, the effect sizes for PTSD-DC, PTSD, and mirror-gazing showed significant evidence of funnel plot asymmetry (see Table 1 and Supplementary Materials for funnel plots). By contrast, significant funnel plot asymmetry was observed for all pharmacological agents except N_2_O and psychedelics. These results are potentially reflective of potential publication bias and suggest that effect size estimates for multiple categories may be inflated.

### Meta-regressions

Meta-regression analyses were undertaken to examine whether variability in effect sizes across studies was attributable to different methodological features across studies including the induction method, administration method, sample, design, and other methodological features. Our first set of meta-regression analyses compared state dissociation effects across categories in cases where *k*≥5 in each category (Supplementary Table 5). Both mirror-gazing and military training elicited larger increases in state dissociation than exposure to trauma and negative affect stimuli (Δ*SMD*s>0.30), but the former two did not significantly differ. By contrast, no significant differences were found across diagnostic categories or pharmacological agents. Baseline dissociation in PTSD-DC was greater than induced dissociation for all induction methods (Δ*SMD*s>0.50) except for mirror-gazing and pharmacological agents. Baseline dissociation in PTSD and MDD was greater than induced effects from trauma and negative affective stimuli exposure but weaker than pharmacological induction effects. Finally, comparisons across categories indicated that induced dissociation was greater in response to ketamine than all psychological techniques (Δ*SMD*s>0.50) whereas the response to esketamine was only greater relative to trauma and negative affective stimuli exposure.

Our final set of meta-regression analyses sought to clarify whether heterogeneity in effect sizes is associated with different methodological features (Supplementary Table 6). Effect sizes were not significantly moderated by methodological quality or administration method (clinician/experimenter vs. self-report). Non-clinical samples displayed a stronger dissociative response to ketamine than clinical samples (Δ*SMD*=0.46), but the two groups did not significantly differ in other categories. Experimental design did not uniformly significantly moderate effect sizes with larger induction effects for between-groups designs and within-groups designs for mirror-gazing and ketamine, respectively. Induction effects did not differ across different types of control conditions for psychological techniques whereas among ketamine studies effect sizes were greater in studies employing inert placebo controls than baseline or active drug controls but were not significantly moderated by dose or route of administration.

## Discussion

This meta-analysis sought to quantify, and compare, baseline state dissociation effects in clinical samples and induced state dissociation effects in response to psychological techniques and pharmacological agents. Baseline state dissociation was elevated in multiple diagnostic categories relative to controls but was most pronounced in individuals with PTSD dissociative and complex subtypes (PTSD-DC). Among induction studies, multiple pharmacological agents elicited pronounced dissociative effects in clinical and non-clinical samples. Mirror-gazing was the most robust psychological technique, closely approximating the dissociative effects of pharmacological agents. These results reinforce state dissociation as a prominent transdiagnostic symptom (2) and demonstrate clinically-significant dissociative states can be reliably induced using a range of methods (9, 10).

Our analyses confirmed the presence of elevated baseline state dissociation across several diagnostic categories. Baseline dissociation was most pronounced in PTD-DC and PTSD, although most studies did not distinguish between PTSD subgroups. Elevated state dissociation in these groups broadly aligns with previous analyses of *trait* dissociation (2), although our results diverge from the latter analysis insofar as individuals with schizophrenia and depressive disorders displayed comparable, albeit weaker, dissociative effects to PTSD in our analysis. Moreover, whereas individuals with FND have been shown to display high levels of trait dissociation, comparable to PTSD (2, 5), FND was characterised by only moderate levels of state dissociation in our analyses and was the only non-significant diagnostic category. This discrepancy is plausibly attributable to a small number of studies including FND samples and the greatest heterogeneity among all diagnostic categories likely driven by differential levels of dissociation in FND subgroups (5).

Although state and trait dissociation are strongly associated, they should be distinguished in research and clinical practice, as state dissociation may indicate more severe psychopathology (25). These results reinforce the importance of measuring dissociation in different diagnostic categories and clinical contexts, particularly given that dissociation may predict broader symptomatology (5–7), and treatment outcomes (26).

Analyses of pharmacological agents revealed that two agents elicited state dissociation effects that were comparable to, or exceeded, baseline dissociation in individuals with PTSD-DC. The most pronounced effects were observed with ketamine and cannabis, with slightly weaker, albeit still large, effects in N_2_O and psychedelics, and esketamine. Further analyses suggested that ketamine’s dissociative effects are greatest the first 50 minutes post-infusion and larger in non-clinical samples. Taken together, these results indicate that different types of pharmacological action can produce large dissociative effects. Accordingly, dissociative states might not be associated with the perturbation of a specific neurochemical system but rather with broader network-level increases in neural signal complexity and changes in intra- and inter-network connectivity that are shared across these agents (27, 28) and potentially with clinical samples (29) (for a consideration of neurophysiological differences across some of these agents, see (30)). For example, ketamine, N_2_O, and LSD are all associated with aberrant functional connectivity in nodes of the default mode and dorsal attention networks (e.g., precuneus and temporoparietal junction) (28), which may parallel atypical precuneus and temporoparietal volume and/or functional connectivity in individuals with high dissociation (31–34). These effects may reflect disruptions in embodiment and multimodal integration that play a central role in experiences of depersonalisation and derealisation or distortions in features of subjective experience subserved by a broader posterior cortical hot zone, which is hypothesised to play a critical role in supporting the subjective contents of consciousness (28). Continued research into these other compounds may also help in advancing research into pharmacotherapeutic agents for dissociative symptomatology; for example, whereas N_2_O acts a partial agonist of opioid receptors (30), preliminary research suggests that opioid antagonists seem to reduce dissociative symptoms (35) (see also (36)).

Among psychological techniques for inducing dissociative states, mirror-gazing was the only method that elicited comparable dissociative effects to those observed in diagnostic categories and with pharmacological agents. In particular, the magnitude of the dissociative response to mirror-gazing was similar to baseline dissociation in PTSD (and larger than all categories except PTSD-DC) and induced dissociation in response to esketamine but weaker than all other pharmacological agents. The neurocognitive substrates of mirror-gazing remain largely unknown but it may produce dissociative states, particularly depersonalisation, through a partial decoupling of visual and cognitive self-referential processing (1, 37). By contrast, stress induction methods used in military/survival training elicited weaker, albeit still large, effects that were larger than moderate and non-significant effects for exposure to trauma stimuli and negative affect stimuli, respectively. The greater efficacy of the former is plausibly attributable to its status as a more uniform stressor than tasks involving different types of stimulus presentation with variable effects across individuals. Techniques targeting awareness and attention (sleep deprivation, complementary methods) also produced moderate dissociative effects, which aligns with accumulating evidence for a link between sleep disturbances and dissociation (1). Although typically viewed as a consequence of stress (1, 3), these results cumulatively indicate that dissociative states can be reliably induced through a variety of methods including by modulating awareness, perception, and sleep and highlight the need for direct comparisons of these methods and their neurocognitive substrates (1, 9).

The observed state dissociation effects have direct implications for the development of an experimental model of dissociation (38). The cumulative data point to the greater efficacy of mirror-gazing relative to stress induction methods, given that it produces larger dissociative effects and is less likely to trigger adverse events (9, 39). Our results additionally highlight ketamine, cannabis, and N_2_O as the most robust pharmacological agents for inducing dissociation; the latter is particularly well-suited to experimental research given that its low blood solubility elicits rapid induction and termination effects (10, 39). Although these results are not formally incompatible with broad consensus that dissociative psychopathology is a consequence of developmental trauma (40), they underscore the need for direct comparisons between methods. Preliminary research suggests that script-driven imagery methods of inducing dissociation seem to be associated with activation patterns (e.g., greater amygdala activation (41)) that differ from those involving pharmacological agents (28). Accordingly, further neurophysiological research comparing different methods is necessary to understand the extent to which these methods have overlapping and distinct neurocognitive substrates. Preliminary trends suggest that different pharmacological agents and mirror-gazing produce greater derealisation than depersonalisation; further targeting these effects could be beneficial in elucidating the neural correlates of subdimensions of dissociation (42). Development of experimental models of dissociation will also require greater attention to the temporal dynamics of, and dosing effects on, state dissociation, which are poorly understood apart from ketamine. Our analyses suggest that clinical samples display weaker dissociative responses to ketamine and previous research points to trait dissociation as a predictor of such responses (26); further attention to the sources of individual differences in response to induction methods is necessary. Finally, although our meta-analysis demonstrates that mirror-gazing and multiple pharmacological agents can induce dissociative states that are large in magnitude and comparable to baseline dissociation in some clinical samples, further research is required to assess their clinical relevance in comparison to dissociative effects in diagnostic categories.

### Limitations

The principal limitations of this meta-analysis concern limited available data in specific categories and methodological weaknesses in the original studies. Many categories included a small number of effect sizes, thereby limiting the precision of our estimates and preventing us from examining sources of heterogeneity. Our choice to restrict our analyses to studies using the CADSS facilitated comparisons across categories and ensured a good degree of phenomenological uniformity in response patterns but may have excluded important research with other validated instruments (16). In turn, it will be important for future empirical studies and meta-analyses to compare and contrast the CADSS with these other measures. Only a small proportion of studies reported CADSS subscale scores (e.g., depersonalisation) thereby limiting our analyses of different subdimensions of state dissociation. It remains unclear whether this omission reflects publication bias, poor psychometric properties of specific subscales, or other factors but further research into these subscales and their psychometric properties and discriminant validity is required. Only a small minority of studies included trait dissociation measures, which prevented us from assessing their value in predicting dissociation induction effects (26). State dissociation was alternately measured *peri*-induction (most pharmacological agents) or post-induction (most psychological techniques), which may introduce different response biases that were not captured in our analyses. Relatedly, most of the original studies are potentially confounded by demand characteristics and potential placebo effects as participants are likely to become unblinded to experimental conditions due to psychoactive effects (43). We planned to probe this in our pre-registered analyses by examining the presence of suggestions for dissociative responses during procedures, but this information was not reliably reported and could not be analysed. Insofar as dissociation was typically measured as a secondary outcome or adverse event (13), these types of biases may be less pronounced than for psychedelic effects but further consideration of this issue is warranted, such as through the use of active drug controls, stringent reporting of suggestion effects, and statistical corrections for unblinding effects (44).

Aside from ketamine, studies did not report state dissociation at multiple time points, thereby disenabling systematic analyses of peak dissociation effects. We were unable to examine the potential confounding effects of concurrent psychotropic medication in clinical samples. Except for ketamine, we were unable to examine the moderating impact of dose on state dissociation effects due to small sample sizes. Moreover, most studies reported ketamine (and other agent) doses as mg/kg, which does not account for individual differences in drug absorption, metabolism, distribution, and excretion (45), leading to variability in plasma concentrations and dissociative effects that could not be captured in our ketamine dose analyses. For this reason, our observation of a non-significant effect of ketamine dose on state dissociation should be treated with caution. Many of the agents we analysed elicit broader psychotomimetic effects (e.g., hallucinations) that could overshadow more subtle dissociative responses (10, 12, 46), thereby potentially limiting the measurement reliability of state dissociation (47).

### Summary and conclusions

This meta-analysis confirmed that state dissociation is a transdiagnostic symptom present in multiple psychiatric conditions that can be reliably induced using different pharmacological agents as well as mirror-gazing. These findings have direct implications for the experimental modelling of dissociation in controlled research, the search for neurophysiological markers of dissociation, and the assessment of adverse events and treatment outcomes in psychopharmacological interventions involving NMDAR antagonists and classic psychedelics.

## Data Availability

All data are available from the original papers.

## Acknowledgements

LW is supported by the Department of Psychology, Institute of Psychiatry, Psychology & Neuroscience, King’s College London. SP is supported by a Medical Research Council Fellowship. SKK is supported by the Medical Research Council (UK). DBT is supported by the Gyllenbergs Foundation.

## Notes

### Competing Interest Statement

The authors have declared no competing interest.

### Author Declarations

The data were openly available prior to conducting the meta-analysis and are available by accessing the original papers.

### Summary of Updates

The analyses have been updated to remove the inclusion of multiple effect sizes from the same samples. The results have not changed substantially from the previous version.

