## Supplementary for "The induction of dissociative states: A meta-analysis"

16 De Crespigny Park

London, UK SE5 8AB

**Supplementary Methods**

*Search terms*

The following search terms were used in the databases:

Clin* Admin* Diss* State* Scale" OR CADSS OR "dissociative state*" OR "state dissoc*" OR SADSS.

**Supplementary Figure 1.** *PRISMA Flowchart of Study Selection Process.*

**Identification of studies via databases**

Records removed *before screening*:

Duplicate records removed (n = 1,669)

Records identified:

Databases (n = 3883)

**Identification**

Records screened

(n = 2214)

Records excluded

(n = 1738)

Reports excluded:

Not journal article (n = 89)

No CADSS (n = 59)

Missing data (n = 57)

Overlapping data (n = 43)

No control condition (n = 42)

Wrong study design (n = 21)

Other reasons (n = 20)*

Reports assessed for eligibility

(n = 476)

**Screening**

Reports assessed for analysis

(n = 145)

Insufficient number of effect sizes in a category

(n = 17)

**Included**

Studies included in review

(n = 128)

*Notes*. *Full list of reasons in Reasons for exclusion.

*Reasons for exclusion*

The ‘Other reasons (20)’ in the PRISMA flow chart (Supplementary Figure 1) are as follows: duplicate (7), adolescent/child population (5), no results presented/available (4), not in English (3), and no peer-review (1). ‘Wrong study design (21)’ contains reports excluded as wrong study design (12), reduction study (6), and longitudinal study (3). ‘Not journal article (89)’ contains reports excluded as abstract (38), supplement (13), review or meta-analysis (10), case study (9), poster (9), post-hoc analysis of previously published results (5), dissertation (2), letter to editor (2), and book chapter (1).

*Methodological quality*

As mentioned in the main text, methodological quality was rated according to 12-15 items concerning study objectives, participant recruitment, demographic data, inclusion and exclusion criteria, clarity of procedure, blinding, pre-registration, and relative matching of groups/conditions.

**Supplementary Table 1.** Methodological quality items.

Endorsement of criteria (1 = criteria met, 0 = criteria not met)

| 1. | Was there a clear specification of study objectives? |
| --- | --- |
| 2. | Was it clearly described where participants were drawn from (e.g. University etc)? |
| 3. | Was it clearly described how participants were recruited (e.g. advertisement, course credits, volunteers etc)? |
| 4. | Were relevant participant characteristics adequately described (age, sex etc)? |
| 5. | Was there a clear description of the inclusion and exclusion criteria? |
| 6. | Was the procedure clearly specified? |
| 7. | Was the experimenter blind to group/condition? |
| 8. | Was the participant blind to condition? (if applicable) |
| 9. | Were complete outcome data (i.e., Ms and SDs) available (e.g. reported in article or given via response to data request)? |
| 10. | Was the study pre-registered? |
| 11. | Were the groups comparable in terms of demographics? (if applicable) |
| 12. | Were the experimental and control conditions relatively matched in terms of potential confounds? (if applicable) |
| 13. | Were the diagnosis procedures and criteria clearly described? (if applicable) |
| 14. | Were the experimental procedures (psychological studies) clearly described? (if applicable) |
| 15. | Were the drug procedure and administration procedures clearly described? (if applicable) |

*State dissociation categories*

Effect sizes were grouped into categories by diagnostic categories, psychological technique, or pharmacological agent. When an insufficient number of effect sizes were available, subcategories were included in higher-order categories including: dissociative and complex subtypes of PTSD; PTSD-DC) (1, 2); major depressive disorder (treatment resistant depression, bipolar disorder, and bipolar depression); functional neurological disorder (including conversion disorder and non-epileptic seizures); trauma stimuli (trauma films/scripts, script-driven imagery, and trauma recall); negative affect stimuli (fear conditioning, subliminal threat cues, fearful faces); military training (survival training; simulated captivity; and combat diver training); complementary methods (hypnosis, imagination, and yoga); cannabis (cannabinoids, tetrahydrocannabinol, and synthetic derivates thereof; e.g., (3, 4); and psychedelics (Ayahusca, 5-methoxy-N,N-dimethyltryptamine [5-MeO-DMT], lysergic acid diethylamide [LSD], and 3,4-methylenedioxymethamphetamine [MDMA]).

**Supplementary Analyses**

The following pre-registered analyses were unable to be performed due to an insufficient number of effect sizes or because the respective variables were unable to be coded due to insufficient information in the original studies: trait dissociation scale scores in different groups; correlations between trait dissociation and CADSS scores; and presence of suggestions for dissociative experiences in response to the induction methods. Similarly, trim-and-fill estimates were not performed because of small sample sizes in most categories and controversy regarding the value of this technique.

**Supplementary Results**

159 effect sizes (corresponding to *n*=128 papers) could be included in our analysis categories (see Supplementary Figure 1). Among these 159, *k*=4 (corresponding to *n*=4 papers) only reported CADSS subscale scores (not total scores) and were only included in the meta-analyses of CADSS subscale scores (see below) and *k*=2 (corresponding to *n*=1 papers). Accordingly, the main analyses were performed on the remaining 155 effect sizes (corresponding to *n*=123 papers), prior to outlier removal (see main text).

The principal reason for exclusion of effect sizes from the main analyses was that there was an insufficient number of effect sizes (*k*<3) in a specific category; effect sizes overlapped across two categories (e.g., administration of a pharmacological agent *and* a psychological technique); effect sizes yielded extreme values (e.g., Infinity) due to low descriptive statistics (*M*=0, *SD*=0) in one condition; or effect sizes corresponded to timepoints that were not specified or were outside the time window of interest (>90 minutes post-infusion; ketamine and esketamine studies only).

**Supplementary Table 2.** *Characteristics of included studies as a function of category.*

| **Category** | |  | |  | | **Gender (*M* %)** | | **Methodological quality rating** | |
| --- | --- | --- | --- | --- | --- | --- | --- | --- | --- |
| **Study Type** | | ***k*** | | ***n(study)**** | | ***% female*** | ***% male*** | ***M*** | ***SD*** |
| **Diagnostic categories** | | | | | | | | | |
| PTSD-DC | | 7 | | 7 | | 76 | 24 | 0.64 | 0.11 |
| PTSD | | 12 | | 12 | | 77 | 23 | 0.64 | 0.07 |
| MDD | | 6 | | 5 | | 54 | 46 | 0.67 | 0.16 |
| SZ | | 3 | | 3 | | 0 | 100 | 0.63 | 0.10 |
| FND | | 4 | | 4 | | 81 | 19 | 0.63 | 0.10 |
| **Psychological techniques** | | | | | | | | | |
| Mirror-gazing | | 12 | | 8 | | 89 | 11 | 0.67 | 0.10 |
| Military training | | 9 | | 8 | | 17 | 83 | 0.55 | 0.11 |
| Sleep deprivation | | 3 | | 3 | | 30 | 70 | 0.56 | 0.05 |
| Trauma stimuli | | 18 | | 15 | | 66 | 34 | 0.62 | 0.13 |
| Complementary methods | | 3 | | 2 | | 67 | 33 | 0.47 | 0.20 |
| Negative affect stimuli | | 5 | | 3 | | 96 | 4 | 0.66 | 0.12 |
| **Pharmacological agents** | | | | | | | | | |
| Ketamine | | 47 | | 43 | | 40 | 60 | 0.68 | 0.17 |
| Cannabis | | 4 | | 4 | | 41 | 59 | 0.78 | 0.10 |
| N_2_O | | 3 | | 3 | | 46 | 54 | 0.45 | 0.08 |
| Psychedelics | | 4 | | 4 | | 47 | 53 | 0.76 | 0.26 |
| Esketamine | | 6 | | 6 | | 50 | 50 | 0.74 | 0.04 |

*Notes.* Methodological quality mean scores (range: 0-1) and standard deviations for study categories (after excluding outliers). Gender (*M* %) = mean proportion of each gender category across studies (only includes studies that reported gender proportions). FND = functional neurological disorder; MDD = major depressive disorder; N2O = nitrous oxide; PTSD = post-traumatic stress disorder; PTSD-DC = dissociative and complex subtypes of PTSD; SZ = schizophrenia. * *n*<*k* indicates that multiple effect sizes were extracted from a study in the respective category (data reflect quality ratings per study rather than per effect size).

**Forest plots of meta-analysis results**

Supplementary Figures 2-14 present forest plots for all categories except for ketamine, mirror-gazing, and PTSD (see Figures 3, 4, 5). Each square marker (and whiskers) reflects *SMD*s (and 95% CIs) of the original study effect sizes whereas the bottom diamond represents the aggregate effect size, indicating non-significant results when crossing zero. Dissociative states were significantly greater in all reference categories relative to controls except in FND (diagnostic categories) and negative affect stimuli exposure studies (psychological techniques).

**Supplementary Figure 2**. *Forest plot of SMDs (with 95% CIs) for the induction of dissociative states (CADSS scores) at baseline in patients with post-traumatic stress disorder (PTSD). Marker sizes reflect study weights with smaller markers denoting smaller weights. SMD=standardised mean difference.*

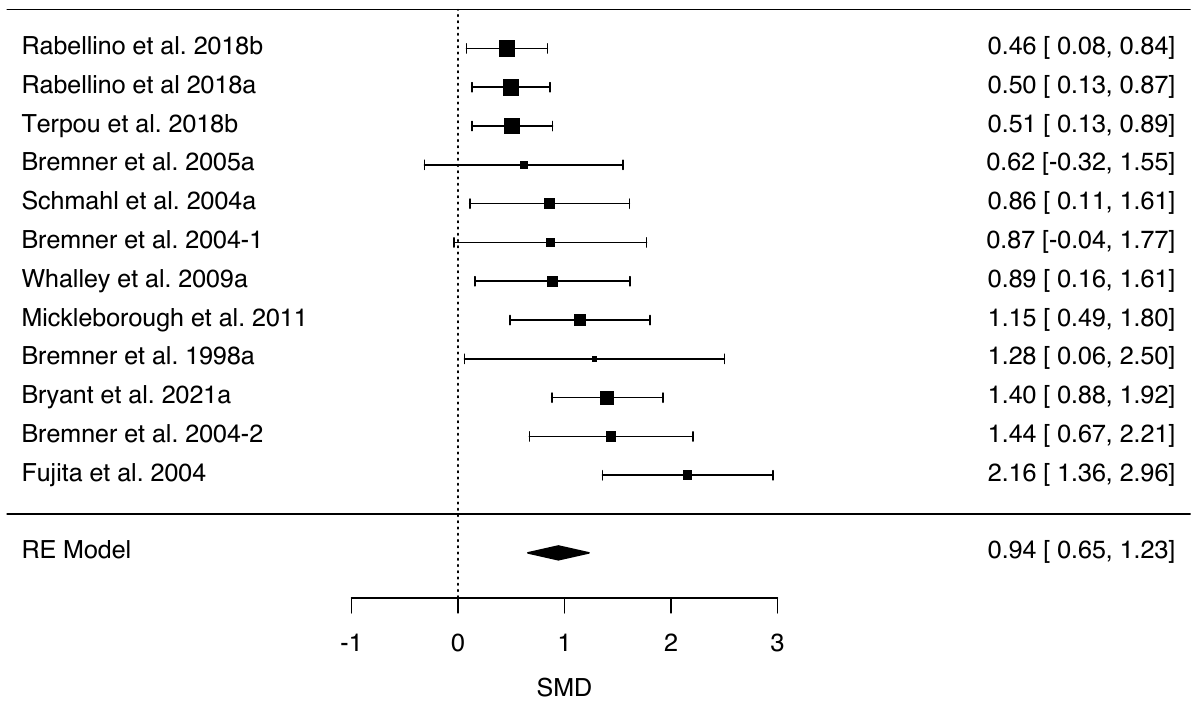

**Supplementary Figure 3**. *Forest plot of SMDs (with 95% CIs) for the induction of dissociative states (CADSS scores) at baseline in patients with schizophrenia. Marker sizes reflect study weights with smaller markers denoting smaller weights. SMD=standardised mean difference.*

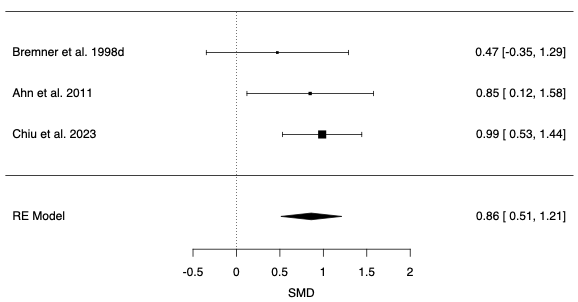

**Supplementary Figure 4**. *Forest plot of SMDs (with 95% CIs) for the induction of dissociative states (CADSS scores) at baseline in patients with major depressive disorder (MDD). Marker sizes reflect study weights with smaller markers denoting smaller weights. SMD=standardised mean difference.*

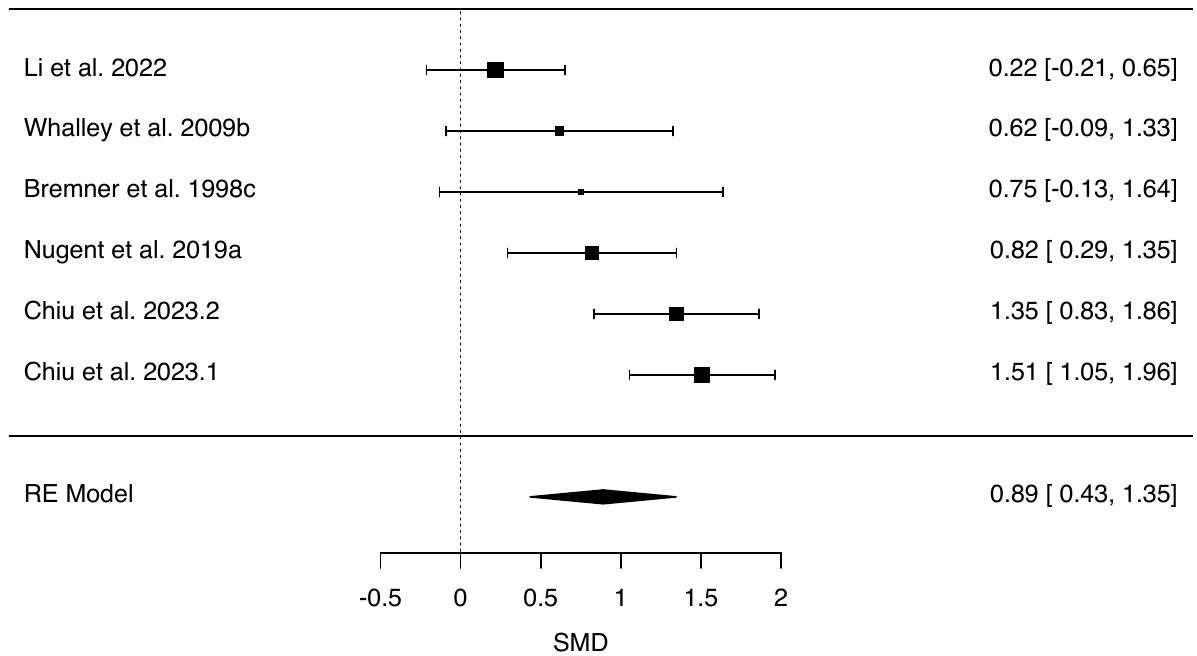

**Supplementary Figure 5**. *Forest plot of SMDs (with 95% CIs) for the induction of dissociative states (CADSS scores) at baseline in patients with functional neurological disorder (FND). Marker sizes reflect study weights with smaller markers denoting smaller weights. SMD=standardised mean difference.*

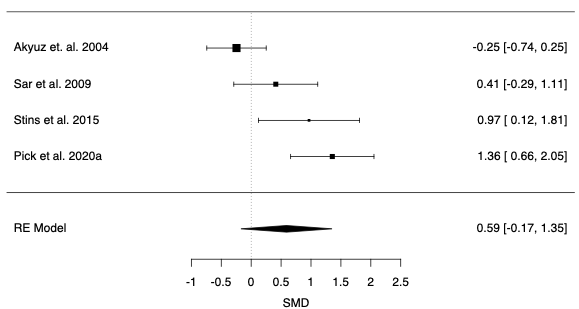

**Supplementary Figure 6**. *Forest plot of SMDs (with 95% CIs) for the induction of dissociative states (CADSS scores) in controlled military training studies. Marker sizes reflect study weights with smaller markers denoting smaller weights. SMD=standardised mean difference.*

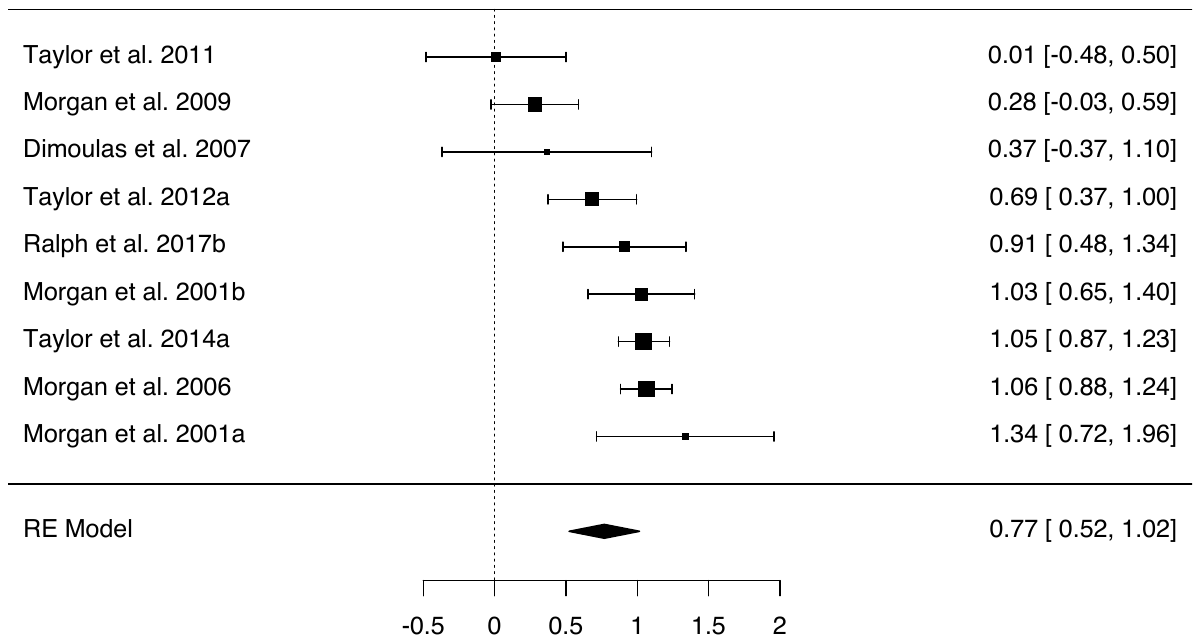

**Supplementary Figure 7**. *Forest plot of SMDs (with 95% CIs) for the induction of dissociative states (CADSS scores) in controlled sleep deprivation studies. Marker sizes reflect study weights with smaller markers denoting smaller weights. SMD=standardised mean difference.*

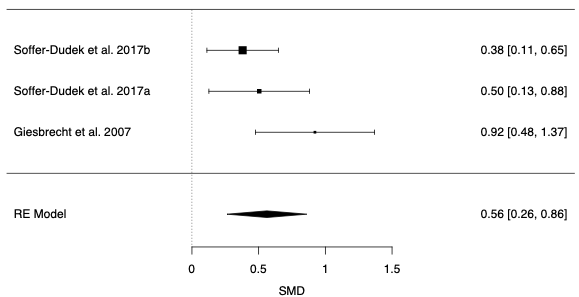

**Supplementary Figure 9**. *Forest plot of SMDs (with 95% CIs) for the induction of dissociative states (CADSS scores) in controlled trauma stimuli studies. Marker sizes reflect study weights with smaller markers denoting smaller weights. SMD=standardised mean difference.*

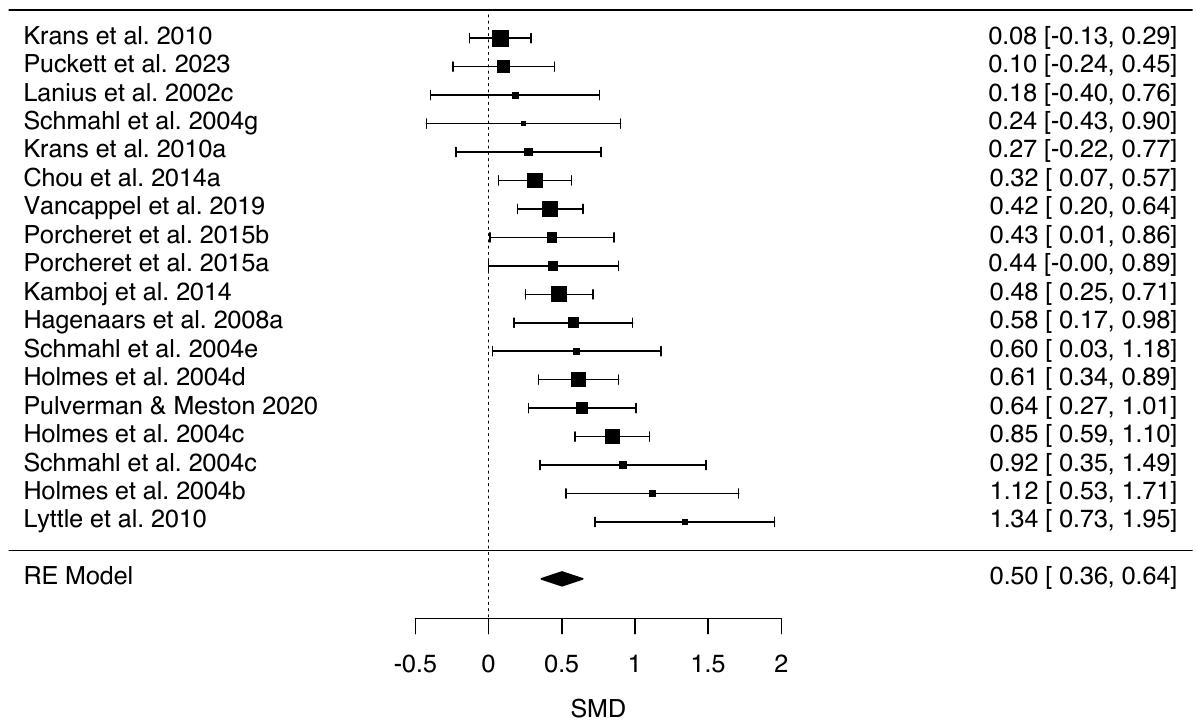

**Supplementary Figure 9**. *Forest plot of SMDs (with 95% CIs) for the induction of dissociative states (CADSS scores) in controlled complementary methods studies. Marker sizes reflect study weights with smaller markers denoting smaller weights. SMD=standardised mean difference.*

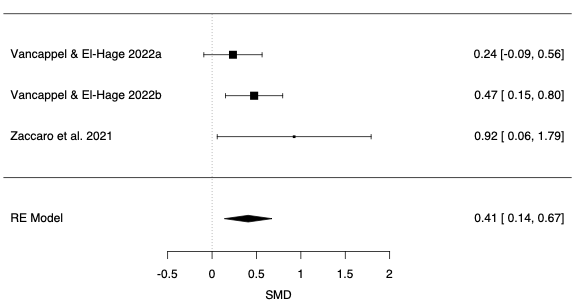

**Supplementary Figure 10**. *Forest plot of SMDs (with 95% CIs) for the induction of dissociative states (CADSS scores) in controlled negative affect stimuli studies. Marker sizes reflect study weights with smaller markers denoting smaller weights. SMD=standardised mean difference.*

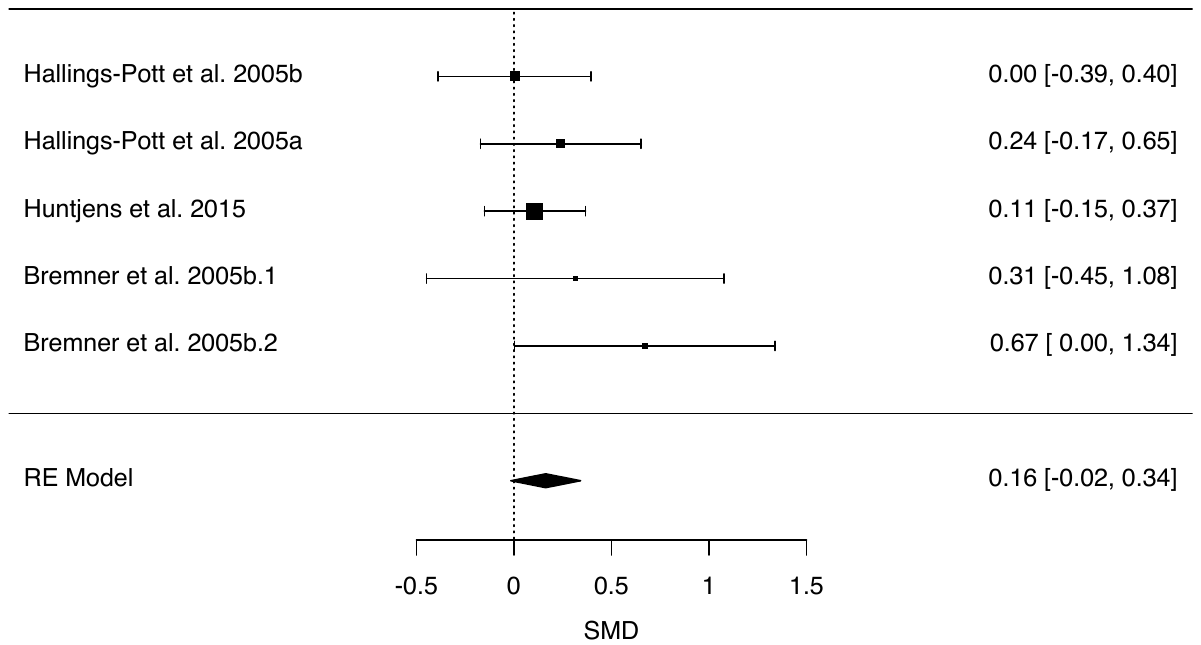

**Supplementary Figure 11**. *Forest plot of SMDs (with 95% CIs) for the induction of dissociative states (CADSS scores) in controlled studies of cannabis administration. Marker sizes reflect study weights with smaller markers denoting smaller weights. SMD=standardised mean difference.*

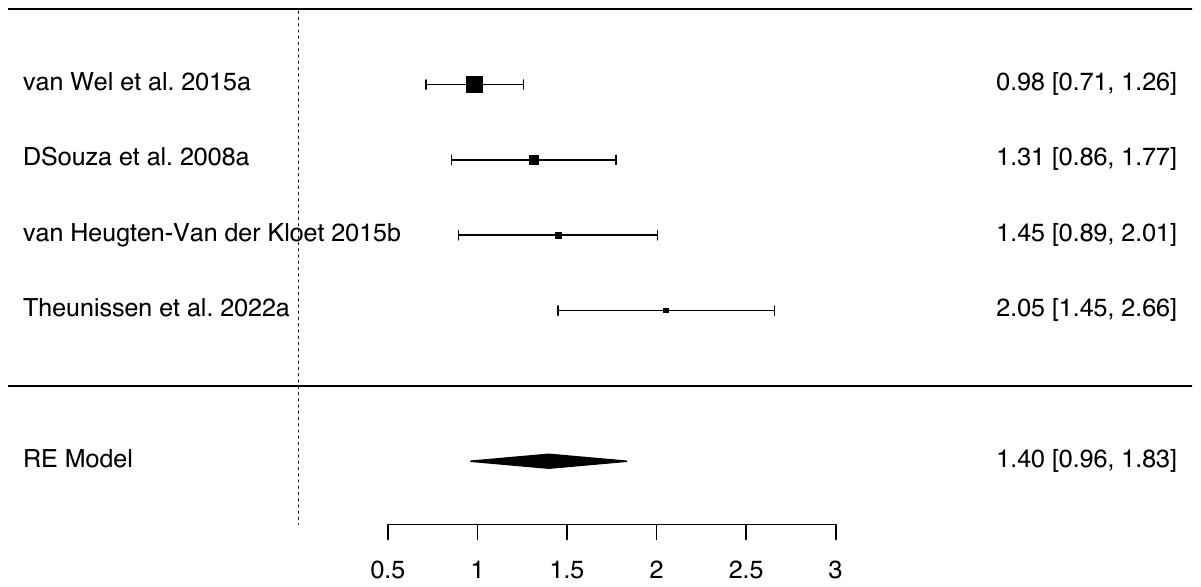

**Supplementary Figure 12**. *Forest plot of SMDs (with 95% CIs) for the induction of dissociative states (CADSS scores) in controlled studies of N2O* *inhalation. Marker sizes reflect study weights with smaller markers denoting smaller weights. SMD=standardised mean difference.*

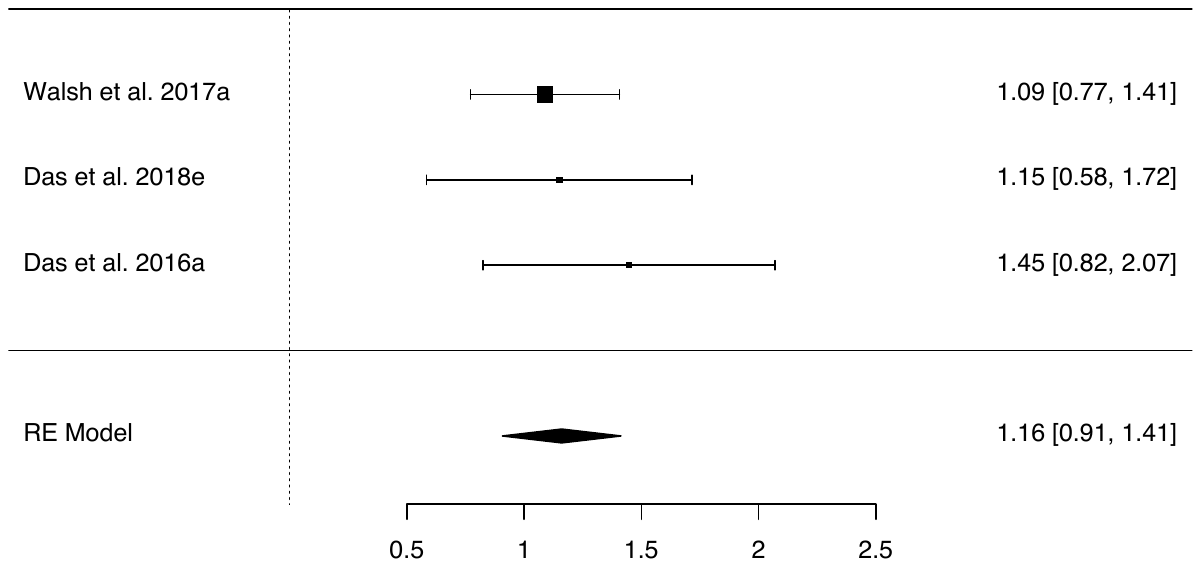

**Supplementary Figure 13**. *Forest plot of SMDs (with 95% CIs) for the induction of dissociative states (CADSS scores) in controlled studies of psychedelics administration. Marker sizes reflect study weights with smaller markers denoting smaller weights. SMD=standardised mean difference.*

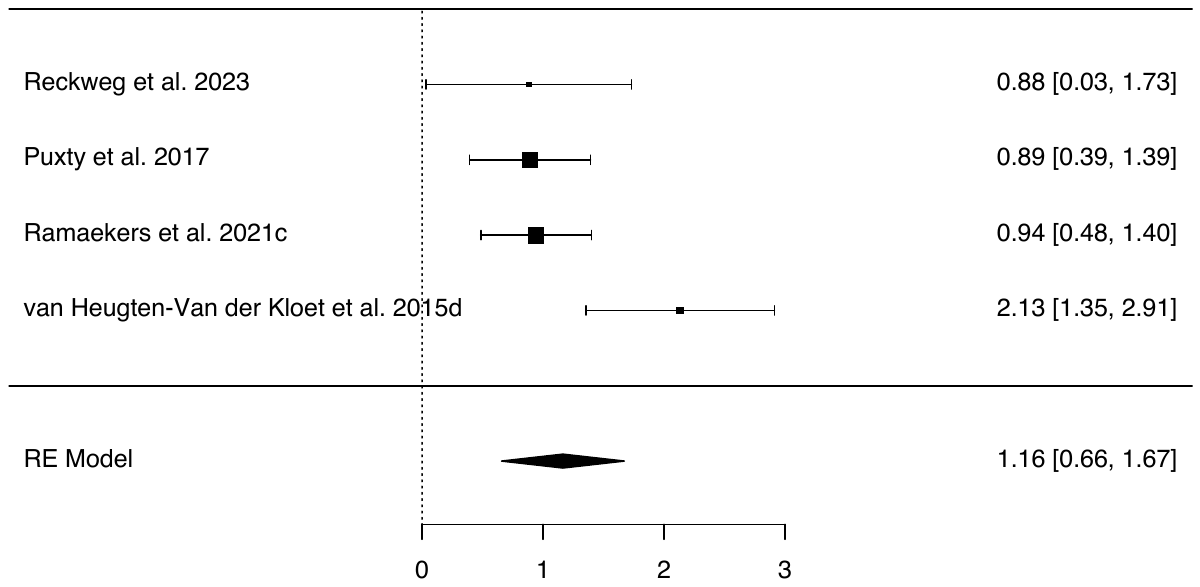

**Supplementary Figure 14**. *Forest plot of SMDs (with 95% CIs) for the induction of dissociative states (CADSS scores) in controlled studies of esketamine administration. Marker sizes reflect study weights with smaller markers denoting smaller weights. SMD=standardised mean difference.*

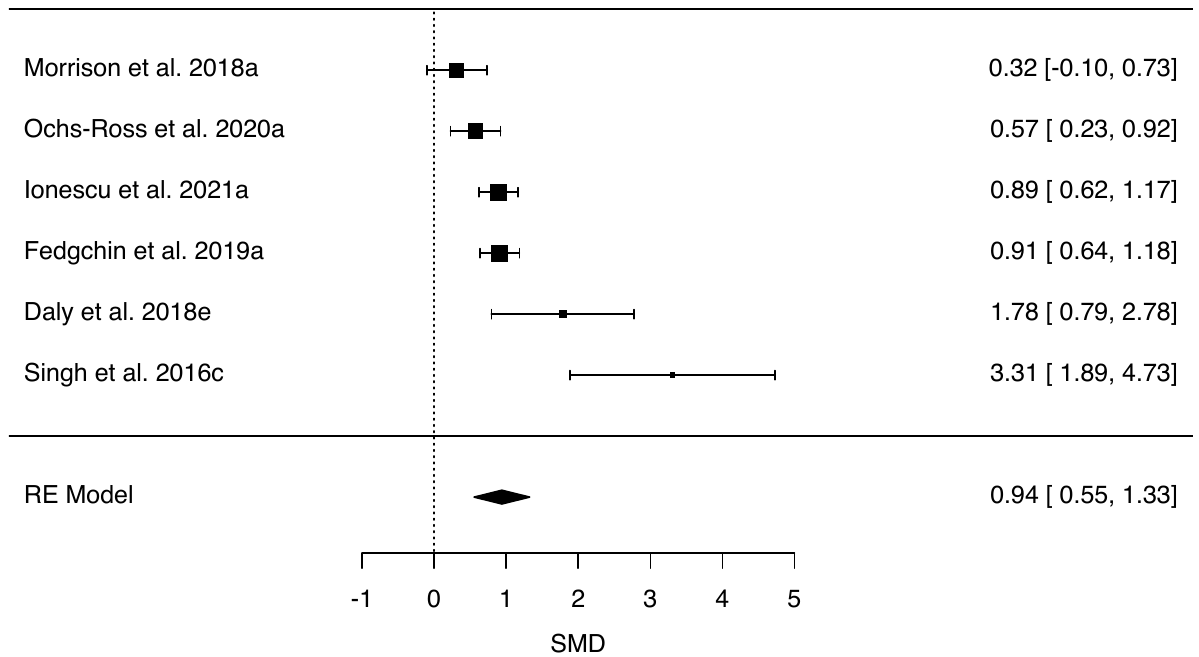

**Publication bias**

Supplementary Figures 15-30 present funnel plots for *SMD*s as a function of *SE*s in all categories. Funnel plot asymmetry is typically interpreted as indicative of publication bias. Categories with small study numbers (*k*s < 10) were included as a reference point but should be interpreted with caution due to poor statistical power. Significant asymmetry (see Table 1) was observed for PTSD-DC, PTSD, mirror-gazing, ketamine, cannabis, and esketamine.

**Supplementary Figure 15.** *Funnel plot of SMDs (with 95% CIs) for the induction of dissociative states (CADSS scores) in controlled studies of post-traumatic stress disorder dissociative and complex subtypes (PTSD-DC).*

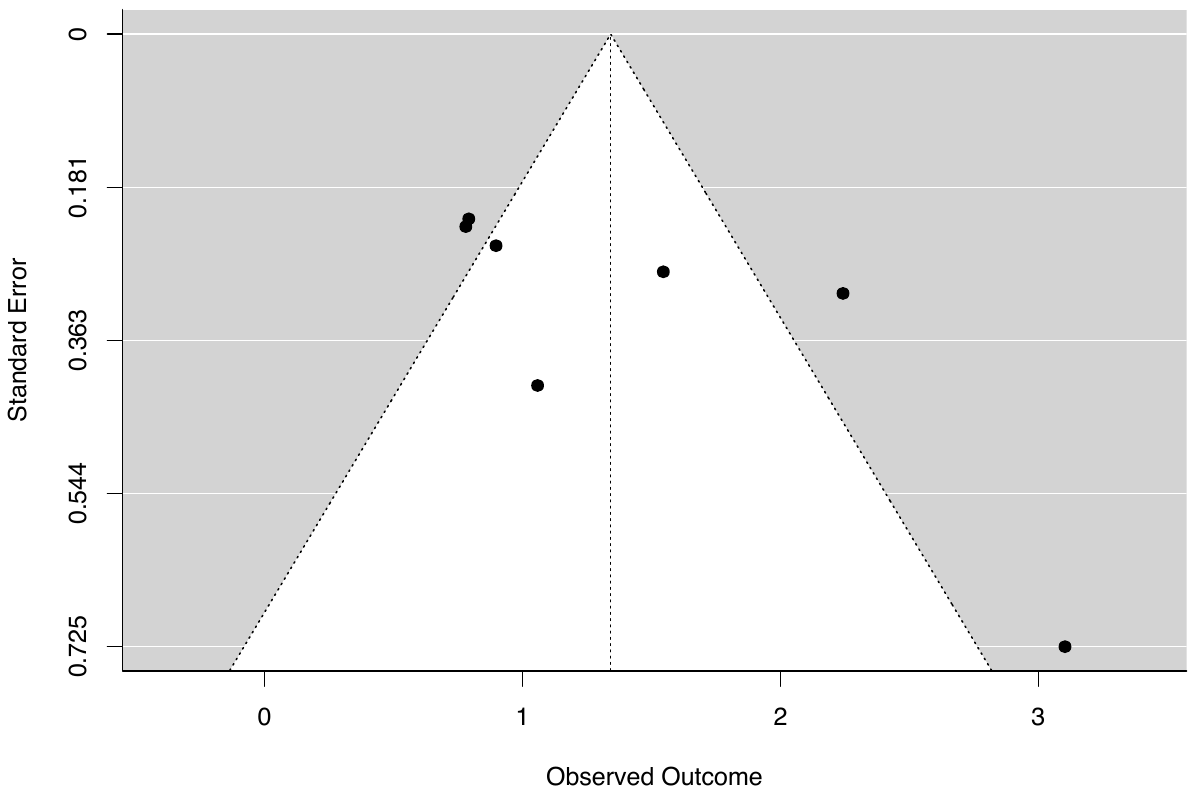

**Supplementary Figure 16.** *Funnel plot of SMDs (with 95% CIs) for the induction of dissociative states (CADSS scores) in controlled studies of post-traumatic stress disorder (PTSD).*

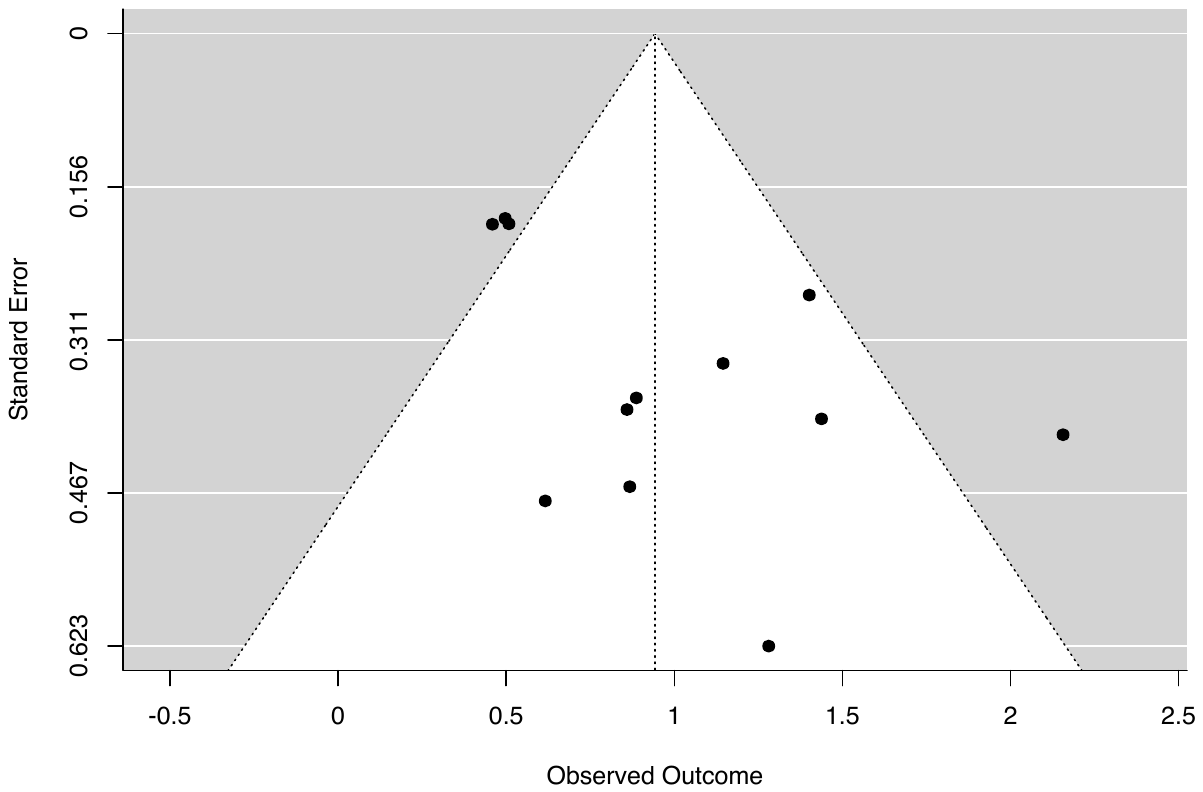

**Supplementary Figure 17.** *Funnel plot of SMDs (with 95% CIs) for the induction of dissociative states (CADSS scores) in controlled studies of major depressive disorder (MDD).*

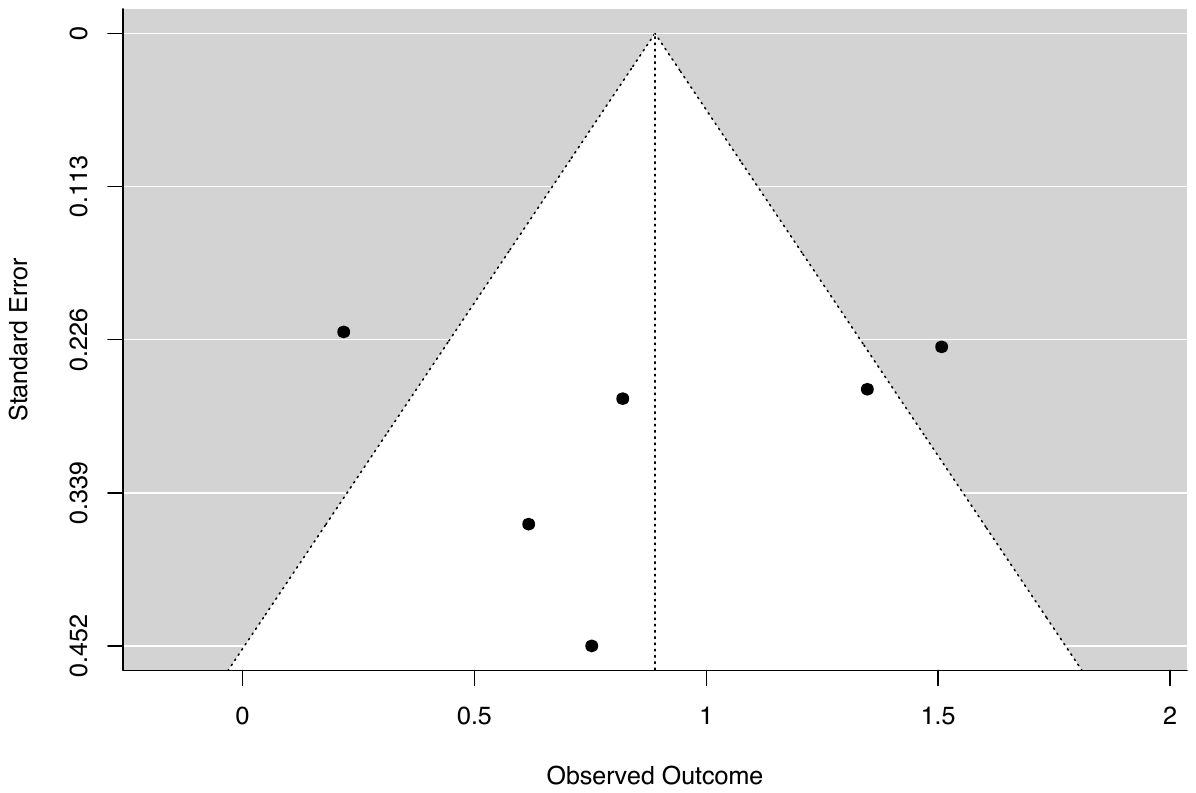

**Supplementary Figure 18.** *Funnel plot of SMDs (with 95% CIs) for the induction of dissociative states (CADSS scores) in controlled studies of schizophrenia.*

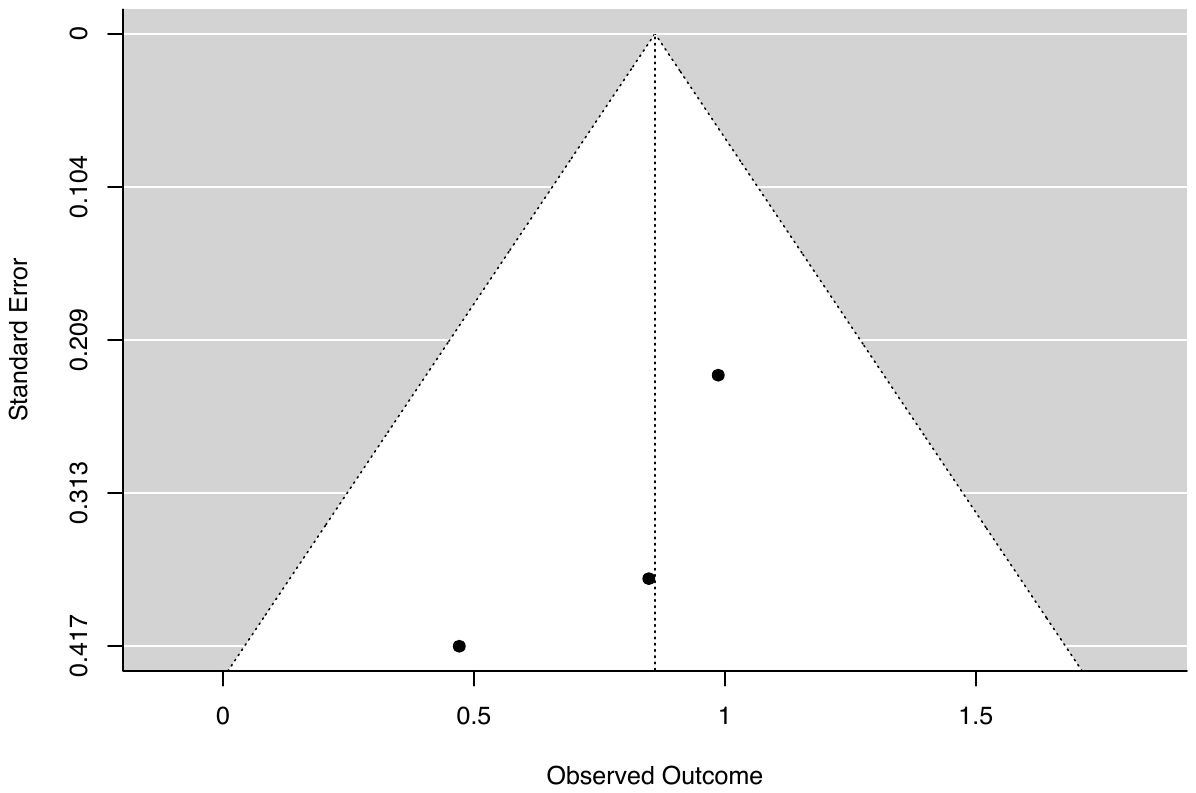

**Supplementary Figure 19.** *Funnel plot of SMDs (with 95% CIs) for the induction of dissociative states (CADSS scores) in controlled studies of functional neurological disorder (FND).*

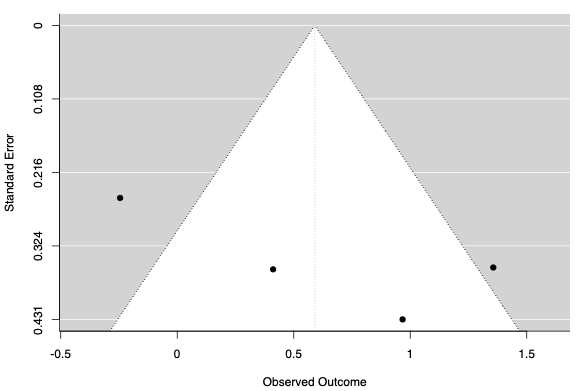

**Supplementary Figure 20**. *Funnel plot of SMDs (with 95% CIs) for the induction of dissociative states (CADSS scores) in controlled studies of mirror-gazing.*

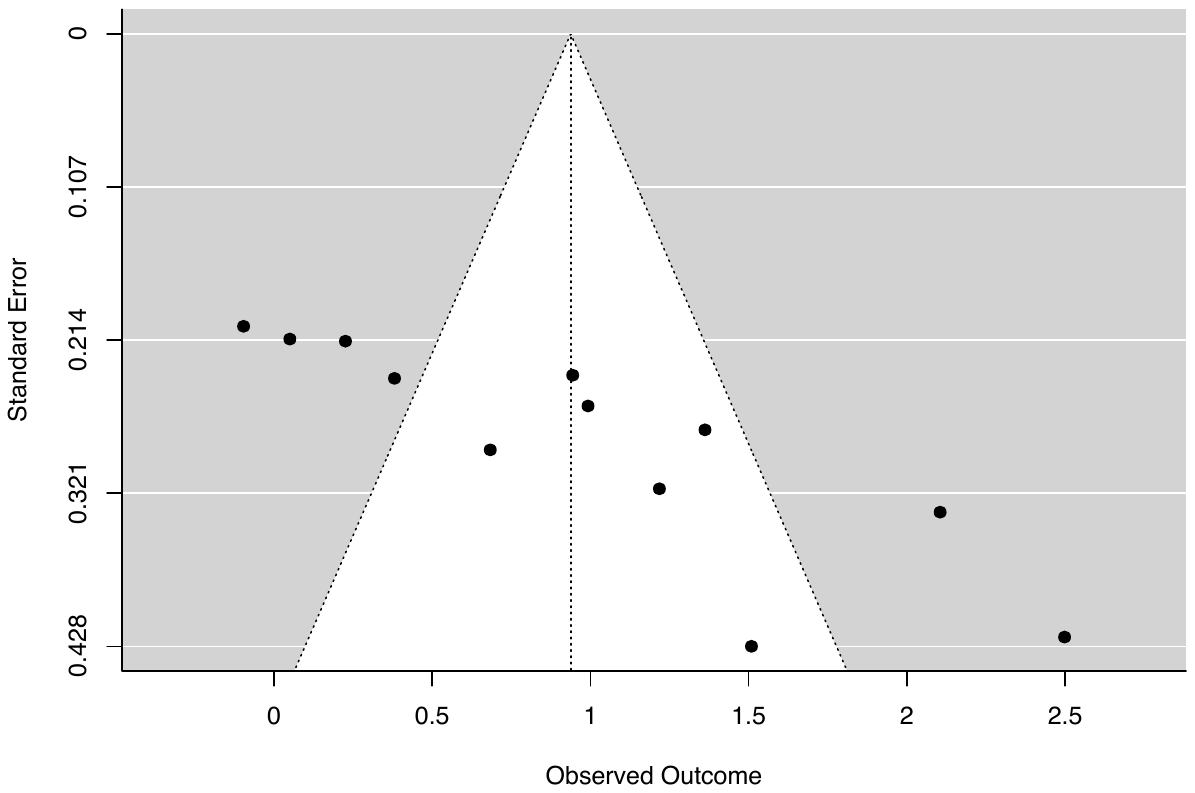

**Supplementary Figure 21.** *Funnel plot of SMDs (with 95% CIs) for the induction of dissociative states (CADSS scores) in controlled studies of military training.*

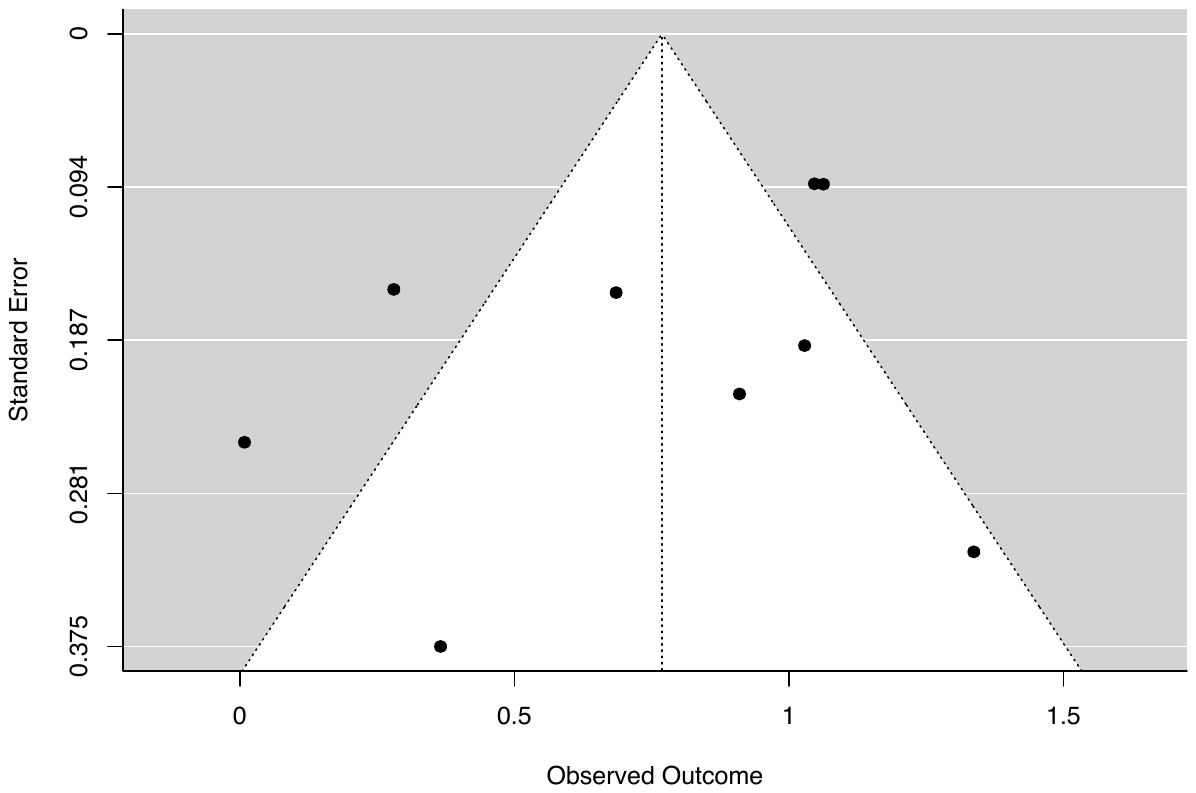

**Supplementary Figure 22.** *Funnel plot of SMDs (with 95% CIs) for the induction of dissociative states (CADSS scores) in controlled studies of sleep deprivation.*

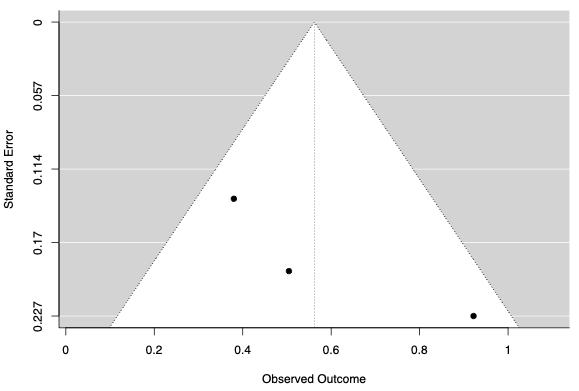

**Supplementary Figure 23.** *Funnel plot of SMDs (with 95% CIs) for the induction of dissociative states (CADSS scores) in controlled studies of trauma stimuli.*

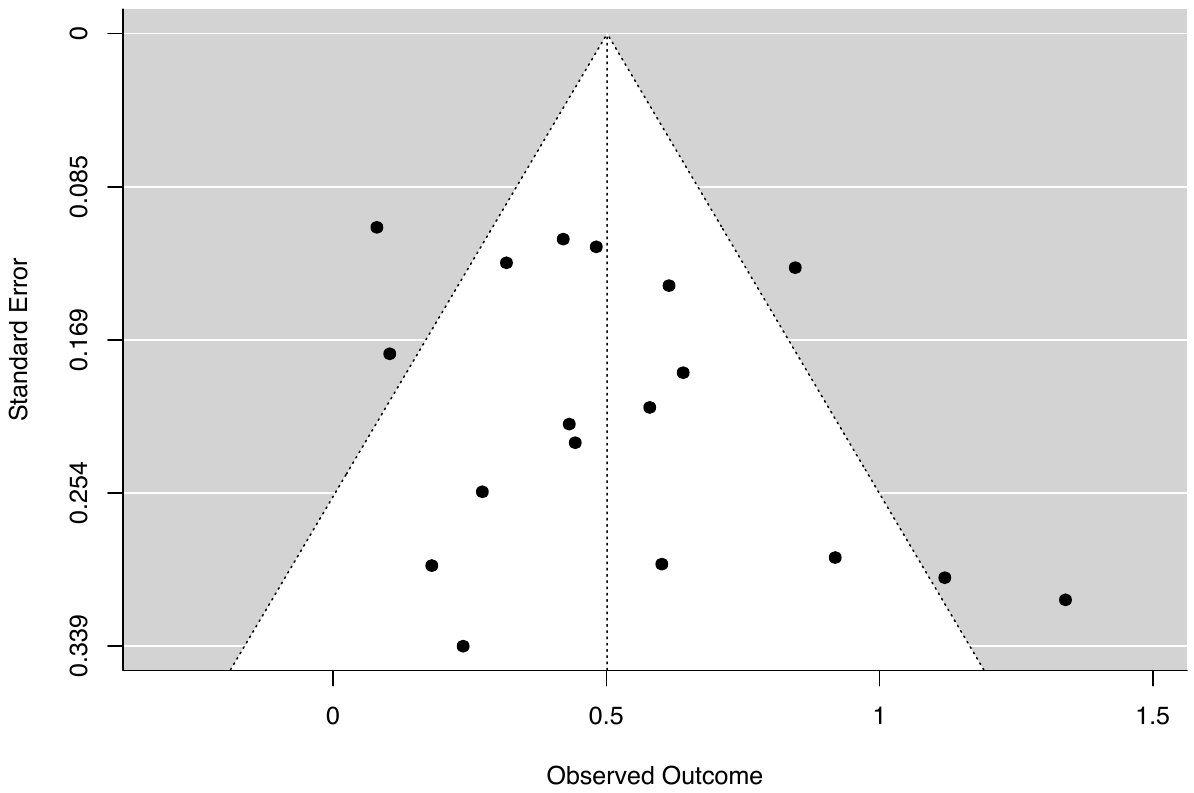

**Supplementary Figure 24.** *Funnel plot of SMDs (with 95% CIs) for the induction of dissociative states (CADSS scores) in controlled studies of complementary methods.*

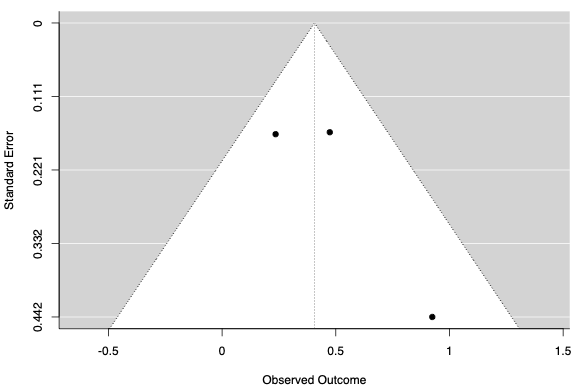

**Supplementary Figure 25.** *Funnel plot of SMDs (with 95% CIs) for the induction of dissociative states (CADSS scores) in controlled studies of negative affect stimuli.*

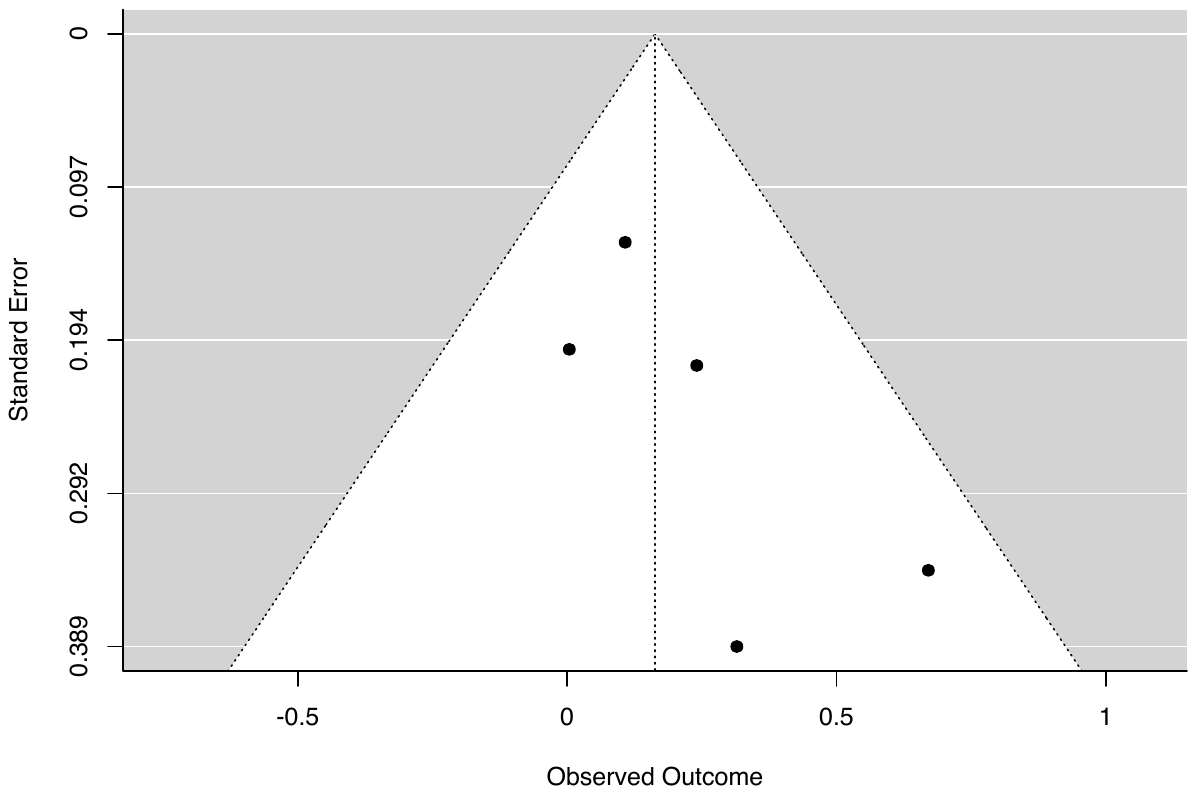

**Supplementary Figure 26.** *Funnel plot of SMDs (with 95% CIs) for the induction of dissociative states (CADSS scores) in controlled studies of ketamine administration.*

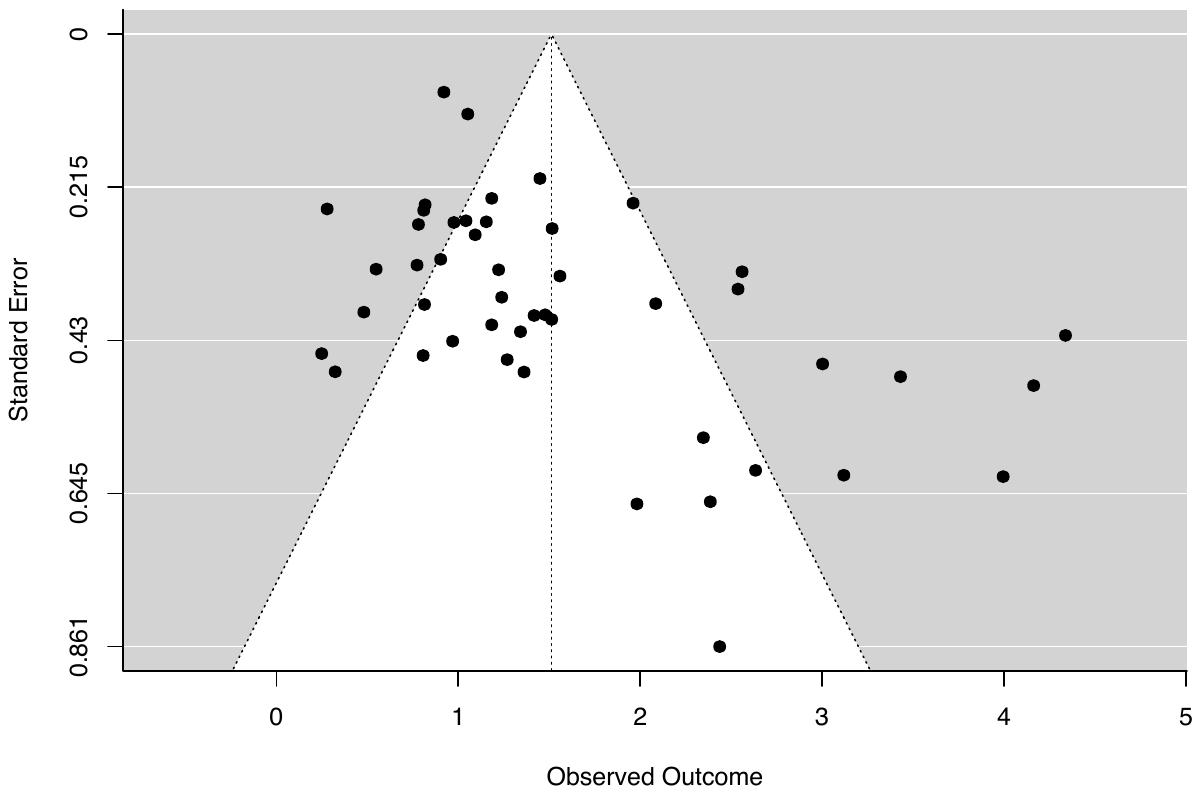

**Supplementary Figure 27.** *Funnel plot of SMDs (with 95% CIs) for the induction of dissociative states (CADSS scores) in controlled studies of cannabis administration.*

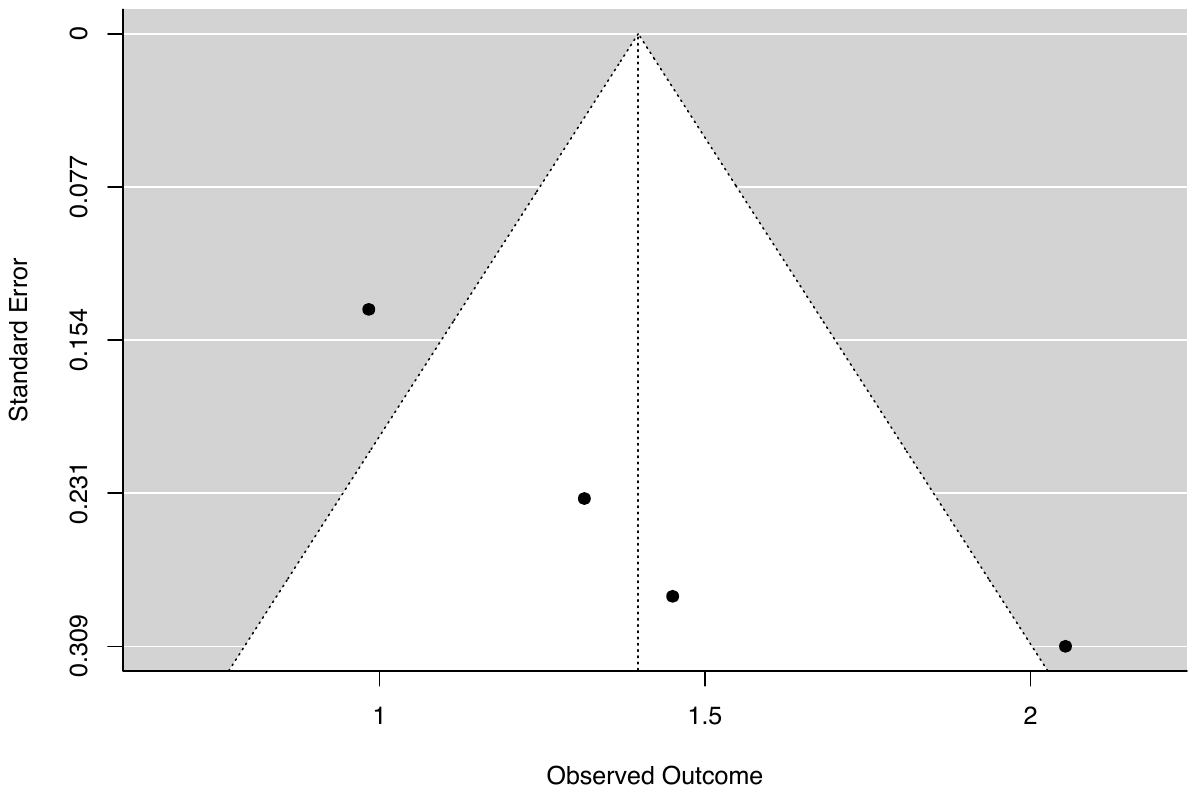

**Supplementary Figure 28.** *Funnel plot of SMDs (with 95% CIs) for the induction of dissociative states (CADSS scores) in controlled studies of N2O inhalation.*

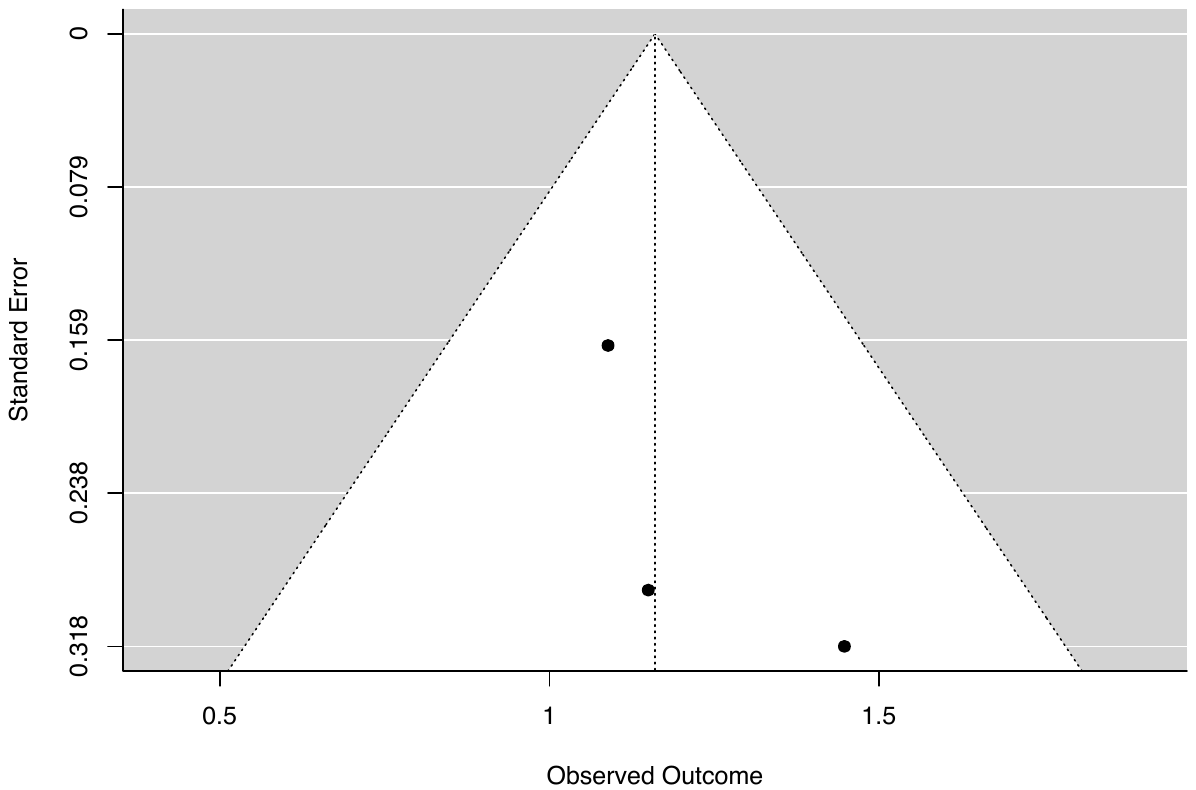

**Supplementary Figure 29.** *Funnel plot of SMDs (with 95% CIs) for the induction of dissociative states (CADSS scores) in controlled studies of psychedelics administration.*

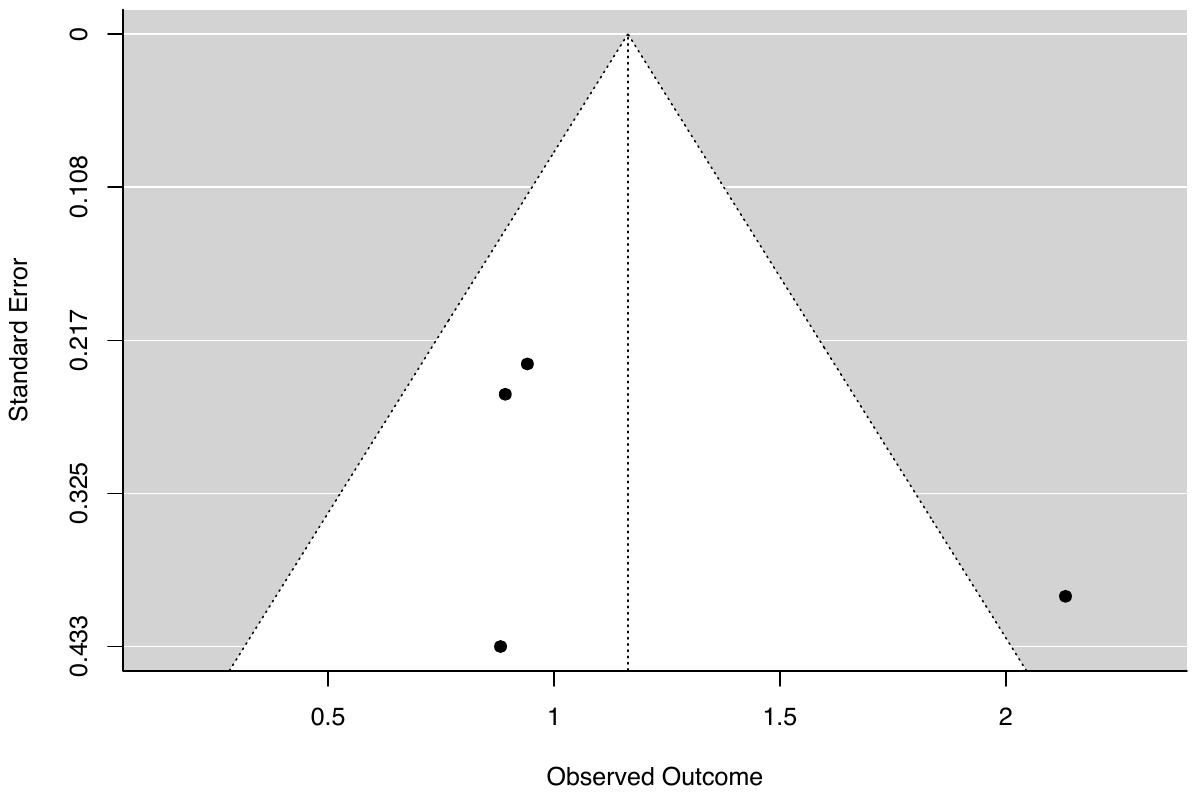

**Supplementary Figure 30.** *Funnel plot of SMDs (with 95% CIs) for the induction of dissociative states (CADSS scores) in controlled studies of esketamine administration.*

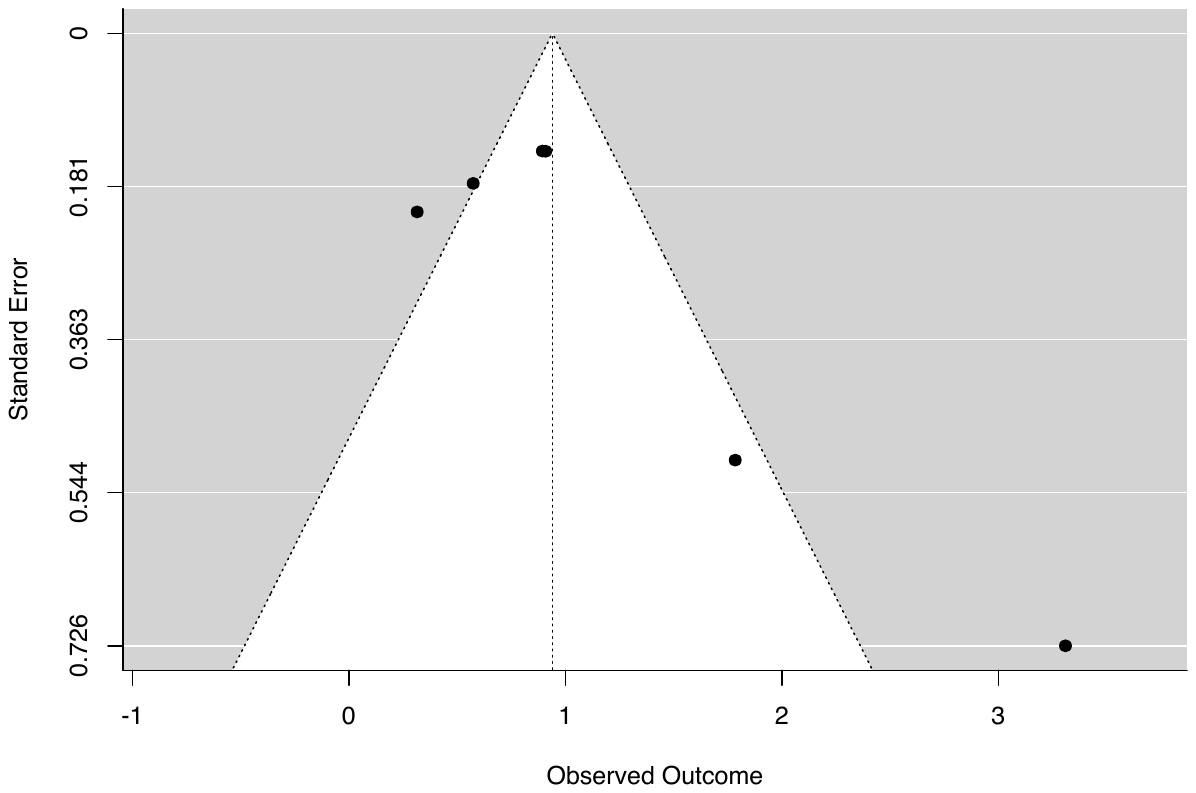

**Meta-analyses of CADSS subscales**

Supplementary Tables 3-4 presents the results of meta-analyses in CADSS subscales and in different subgroups.

**Supplementary Table 3.** Results of meta-analyses of state dissociation subdimensions (CADSS subscale scores) as a function of induction category.

| **Category** | ***k*** | ***N*** | ***SMD*** | **95% CI** | ***PIs*** | ***Z*** | ***p*** | *I*^2^ **(%)** | *T*^2^ | **FPA *p*** | **Outliers** |
| --- | --- | --- | --- | --- | --- | --- | --- | --- | --- | --- | --- |
| **CADSS: Depersonalization** | | |  |  |  |  |  |  |  |  |  |
| Ketamine | 5 | 54 | 2.05 | [0.83, 3.27] | [-2.50, 6.60] | 3.30 | <.001 | 87.42 | 1.65 | .16 | 1 |
| Cannabis | 3 | 111 | 1.12 | [0.56, 1.69] | [-5.71, 7.95] | 3.90 | <.001 | 80.40 | .20 | .002 | 0 |
| Psychedelics | 4 | 68 | 0.84 | [0.32, 1.36] | [-1.33, 3.01] | 3.17 | .002 | 68.50 | .18 | .005 | 0 |
| Mirror-gazing | 3 | 99 | 0.99 | [-0.06, 2.03] | [-11.90, 13.88] | 1.85 | .064 | 87.57 | .74 | .48 | 0 |
| **CADSS: Derealization** | | |  |  |  |  |  |  |  |  |  |
| Ketamine | 5 | 54 | 2.03 | [0.98, 3.09] | [-1.83, 5.89] | 3.77 | <.001 | 82.74 | 1.17 | .50 | 1 |
| Cannabis | 3 | 111 | 1.48 | [0.88, 2.09] | [-5.84, 8.80] | 4.79 | <.001 | 79.77 | .23 | .06 | 0 |
| Psychedelics | 4 | 68 | 1.14 | [0.56, 1.72] | [-1.34, 3.62] | 3.87 | <.001 | 71.62 | .24 | .38 | 0 |
| Mirror-gazing | 3 | 99 | 1.15 | [0.80, 1.51] | [-1.26, 3.56] | 6.35 | <.001 | 0 | 0 | .83 | 0 |
| **CADSS: Amnesia** |  |  |  |  |  |  |  |  |  |  |  |
| Ketamine | 5 | 54 | 1.86 | [0.75, 2.98] | [-2.28, 6.00] | 3.27 | .001 | 86.18 | 1.36 | .29 | 1 |
| Cannabis | 3 | 111 | 1.02 | [0.43, 1.62] | [-6.26, 8.30] | 3.37 | <.001 | 83.15 | .23 | <.001 | 0 |
| Psychedelics | 4 | 68 | 0.77 | [0.50, 1.03] | [0.18, 1.36] | 5.64 | <.001 | 0 | 0 | .71 | 0 |
| Mirror-gazing | 3 | 99 | 0.46 | [-0.51, 1.44] | [-11.64, 12.56] | 0.93 | .35 | 87.79 | .65 | .18 | 0 |

*Notes. k*=number of included effect sizes; *N*=sample size; *SMD*=standardized mean difference; *PI*s=prediction intervals; *I*^2^=heterogeneity statistic; *T*^2^=heterogeneity statistic; FPA*p*=funnel plot asymmetry *p*-value; outliers=number of outliers removed; CADSS=clinician-administered dissociative states scale.

**Supplementary Table 4.** Results of meta-analyses of state dissociation effects (CADSS score differences; category - control) in response to ketamine infusion at different timepoints of CADSS administration.

| **Category** | ***k*** | ***SMD*** | **95% CI** | ***PIs*** | ***Z*** | ***p*** | *I*^2^ **(%)** | *T*^2^ | **FPA *p*** | **Outliers** |
| --- | --- | --- | --- | --- | --- | --- | --- | --- | --- | --- |
| **Timepoint** | | | |  |  |  |  |  |  |  |
| 5-30 mins | 15 | 1.72 | [1.30, 2.14] | [0.12, 3.32] | 8.06 | <.001 | 83.28 | .50 | .004 | 1 |
| 40-50 mins | 26 | 1.59 | [1.24, 1.93] | [-0.10, 3.28] | 9.06 | <.001 | 88.02 | .64 | <.001 | 3 |
| 60-70 mins | 13 | 1.16 | [0.78, 1.55] | [-0.22, 2.54] | 5.93 | <.001 | 72.97 | .35 | .50 | 1 |
| 75-90 mins | 13 | 0.63 | [0.21, 1.04] | [-1.01, 2.27] | 2.96 | .003 | 89.10 | .51 | .13 | 1 |

**Meta-regression analyses**

Supplementary Tables 5-6 present meta-regression analyses across categories (*k*>4 in each category; Table 5) and within categories (*k*>9 in category; Table 6).

**Supplementary Table 5.** Meta-regression analyses involving category comparisons for state dissociation effects (CADSS score differences; category - control).

| **Comparisons** | ***k*** | **Δ*SMD*** | **95% CI** | ***Z*** | ***p*** | *I*^2^ **(%)** | *T*^2^ |
| --- | --- | --- | --- | --- | --- | --- | --- |
| **Diagnostic categories** |  |  |  |  |  |  |  |
| PTSD-DC v. PTSD | 19 | -0.36 | [-0.88, 0.17] | -1.33 | .18 | 70.15 | .20 |
| PTSD-DC v. MDD | 13 | -0.45 | [-1.11, 0.22] | -1.31 | .19 | 77.31 | .28 |
| PTSD v. MDD | 18 | -0.06 | [-0.57, 0.45] | -0.23 | .82 | 67.29 | .17 |
| **Psychological techniques** |  |  |  |  |  |  |  |
| Mirror-gazing v. military training | 21 | -0.14 | [-0.60, 0.31] | -0.61 | .54 | 84.33 | .22 |
| Mirror-gazing v. trauma stimuli | 30 | -0.36 | [-0.69, -0.03] | -2.11 | .035 | 78.10 | .14 |
| Mirror-gazing v. negative affect stimuli | 17 | -0.67 | [-1.30, -0.04] | -2.09 | .037 | 82.23 | .29 |
| Military training v. trauma stimuli | 27 | -0.27 | [-0.54, -0.003] | -1.98 | .048 | 71.26 | .07 |
| Military training v. negative affect stimuli | 14 | -0.56 | [-0.95, -0.16] | -2.75 | .006 | 72.42 | .08 |
| Trauma stimuli v. negative affect stimuli | 23 | -0.30 | [-0.60, 0.01] | -1.88 | .060 | 58.10 | .05 |
| **Pharmacological agents** |  |  |  |  |  |  |  |
| Ketamine v. esketamine | 53 | -0.45 | [-1.05, 0.14] | -1.51 | .13 | 83.28 | .38 |
| **Psychological techniques v. diagnostic categories** | | |  |  |  |  |  |
| Mirror-gazing v. PTSD-DC | 19 | 0.43 | [-0.24, 1.09] | 1.25 | .21 | 84.45 | .41 |
| Military training v. PTSD-DC | 16 | 0.53 | [0.07, 0.98] | 2.24 | .025 | 79.55 | .14 |
| Trauma stimuli v. PTSD-DC | 25 | 0.75 | [0.40, 1.10] | 4.16 | <.001 | 69.36 | .08 |
| Negative affect stimuli v. PTSD-DC | 12 | 1.06 | [0.52, 1.60] | 3.86 | <.001 | 68.34 | .14 |
| Mirror-gazing v. PTSD | 24 | 0.05 | [-0.47, 0.57] | 0.19 | .85 | 79.96 | .31 |
| Military training v. PTSD | 21 | 0.10 | [-0.31, 0.51] | 0.48 | .63 | 0 | 0 |
| Trauma stimuli v. PTSD | 30 | 0.40 | [0.12, 0.69] | 2.78 | .005 | 62.91 | .07 |
| Mirror-gazing v. MDD | 18 | -0.05 | [-0.72, 0.63] | -0.13 | .90 | 84.20 | .39 |
| Military training v. MDD | 15 | 0.13 | [-0.33, 0.58] | 0.55 | .58 | 78.48 | .12 |
| Trauma stimuli v. MDD | 24 | 0.39 | [0.04, 0.74 | 2.18 | .029 | 67.18 | .07 |
| **Pharmacological agents v. diagnostic categories** | | | |  |  |  |  |
| Ketamine v. PTSD-DC | 54 | -0.15 | [-0.73, 0.43] | -0.51 | .61 | 83.20 | .41 |
| Esketamine v. PTSD-DC | 13 | 0.34 | [-0.27, 0.95] | 1.09 | .27 | 78.82 | .21 |
| Ketamine v. PTSD | 59 | -0.53 | [-0.98, 0.08] | -2.30 | .022 | 81.61 | .37 |
| Esketamine v. PTSD | 18 | 0.01 | [-0.47, 0.49] | 0.04 | .97 | 69.44 | .15 |
| Ketamine v. MDD | 53 | -0.62 | [-1.23, -0.02] | -2.03 | .042 | 83.09 | .40 |
| Esketamine v. MDD | 12 | -0.07 | [-0.65, 0.52] | -0.23 | .82 | 77.21 | .18 |
| **Psychological techniques v. pharmacological agents** | | | |  |  |  |  |
| Mirror-gazing v. Ketamine | 59 | 0.58 | [0.12, 1.04] | 2.46 | .014 | 84.27 | .43 |
| Military training v. Ketamine | 56 | 0.74 | [0.30, 1.17] | 3.33 | <.001 | 83.20 | .31 |
| Trauma stimuli v. Ketamine | 65 | 0.95 | [0.65, 1.26] | 6.16 | <.001 | 80.80 | .24 |
| Negative affect stimuli v. Ketamine | 52 | 1.25 | [0.64, 1.87] | 4.00 | <.001 | 82.43 | .37 |
| Mirror-gazing v. Esketamine | 18 | 0.11 | [0.52, 0.74] | 0.34 | .74 | 84.72 | .32 |
| Military training v. Esketamine | 15 | 0.14 | [-0.29, 0.58] | 0.65 | .52 | 79.64 | .12 |
| Trauma stimuli v. Esketamine | 24 | 0.36 | [0.03, 0.69] | 2.14 | .032 | 68.78 | .07 |
| Negative affect stimuli v. Esketamine | 11 | 0.67 | [0.20, 1.13] | 2.82 | .005 | 66.65 | .09 |

*Notes.* Negative Δ*SMD* values indicate that effect sizes (*SMDs*) for state dissociation differences (category - control) were greater in the first category.

**Supplementary Table 6.** Meta-regression analyses involving comparisons of state dissociation effects (CADSS score differences; category - control) across different methodological features.

| **Comparisons** | ***k*** | **Δ*SMD*** | **95% CI** | ***Z*** | ***p*** | *I*^2^ **(%)** | *T*^2^ |
| --- | --- | --- | --- | --- | --- | --- | --- |
| **CADSS administration (clinician/experimenter v. self-report)** | | | | |  |  |  |
| PTSD | 12 | -0.07 | [-0.99, 0.85] | -0.15 | .88 | 64.77 | .16 |
| Ketamine | 24 | 0.12 | [-0.75, 1.00] | 0.28 | .78 | 82.21 | .43 |
| Mirror-gazing | 12 | 0.31 | [-0.84, 1.47] | 0.53 | .60 | 87.66 | .51 |
| Trauma stimuli | 18 | -0.09 | [-0.41, 0.24] | -0.52 | .61 | 65.60 | .06 |
| **Sample (non-clinical v. clinical)** |  |  |  |  |  |  |  |
| Ketamine | 47 | -0.46 | [-0.90, -0.03] | -2.08 | .038 | 82.64 | 0.40 |
| Mirror-gazing | 12 | -0.06 | [-1.26, 1.13] | -0.11 | .92 | 87.66 | 0.51 |
| Trauma stimuli | 18 | 0.13 | [-0.26, 0.51] | 0.64 | .52 | 65.52 | 0.06 |
| **Design (between v. within-groups)** | |  |  |  |  |  |  |
| Ketamine | 47 | 0.67 | [0.22, 1.12] | 2.90 | .004 | 83.94 | 0.44 |
| Mirror-gazing | 12 | -1.03 | [-1.72, -0.33] | 2.89 | .004 | 79.49 | 0.27 |
| Trauma stimuli | 18 | -0.10 | [-0.50, 0.30] | -0.49 | .63 | 65.72 | 0.06 |
| **Methodological quality** |  |  |  |  |  |  |  |
| PTSD | 12 | 1.53 | [-2.74, 5.80] | 0.70 | .48 | 63.99 | 0.17 |
| Mirror-gazing | 12 | -2.54 | [-7.82, 2.75] | -0.94 | .35 | 86.96 | 0.46 |
| Trauma stimuli | 18 | 0.40 | [-0.92, 1.72] | 0.60 | .55 | 64.85 | 0.06 |
| Ketamine | 47 | 0 | [-0.003, 0.002] | -0.18 | .85 | 84.00 | .43 |
| **Control comparisons** |  |  |  |  |  |  |  |
| Ketamine: baseline v. placebo | 36 | 0.72 | [0.23, 1.21] | 2.86 | .004 | 84.48 | 0.42 |
| Ketamine: baseline v. active drug | 26 | -0.11 | [-0.48, 0.26] | -0.58 | .56 | 58.47 | 0.09 |
| Ketamine: placebo v. active drug | 32 | -0.81 | [-1.53, -0.10] | -2.24 | .025 | 84.11 | .76 |
| Mirror-gazing: group v. condition | 12 | -0.08 | [-0.49, 0.65] | 0.29 | .78 | 87.66 | 0.51 |
| Trauma stimuli: baseline v. control | 18 | 0.02 | [-0.17, 0.21] | 0.20 | .84 | 65.80 | 0.06 |
| **Ketamine administration features** |  |  |  |  |  |  |  |
| Dose (mg/kg) | 39 | -0.16 | [-1.05, 0.74] | -0.34 | .75 | 85.74 | 0.48 |
| Bolus v. infusion | 47 | 0.47 | [-0.47, 1.41] | 0.98 | .33 | 83.56 | 0.42 |

*Notes.* Negative Δ*SMD* values indicate that effect sizes (*SMDs*) for state dissociation differences (category - control) were greater in the first category.
